# Circulating tumor DNA concentration at diagnosis is a modifiable prognostic factor for distant metastatic recurrence in patients with high-risk breast cancer receiving neoadjuvant therapy

**DOI:** 10.64898/2026.07.28.26358343

**Authors:** Mark Jesus M. Magbanua, Denise M. Wolf, Christina Yau, Nayelis A. Manon, Rosalyn W. Sayaman, Lamorna Brown Swigart, Gillian Hirst, Wen Li, Claudine Isaacs, Rebecca Shatsky, Amy S. Clark, Alexandra Zimmer, Rita Mukhtar, Amy L. Delson, Jane Perlmutter, Paula R. Pohlmann, Nola M. Hylton, Rita Nanda, Douglas Yee, W. Fraser Symmans, Laura J. Esserman, Hope S. Rugo, Angela DeMichele, Laura J. van ’t Veer

## Abstract

**Background:** Circulating tumor DNA (ctDNA) is an emerging biomarker of treatment response and recurrence risk, while residual cancer burden (RCB) after neoadjuvant treatment (NAT) is a well-established risk factor for distant recurrence. Here, we examined the association between high ctDNA concentration at diagnosis and risk of distant recurrence after neoadjuvant treatment (NAT), in the context of RCB.

**Methods:** The study included 712 patients with high-risk breast cancer in the neoadjuvant I-SPY2 trial. Tumor-informed ctDNA test results at diagnosis were used to stratify patients into ctDNA-negative and ctDNA-positive groups. For this analysis, the ctDNA-positive group was divided into tertiles (low, intermediate, high) based on ctDNA concentration reported as mean tumor molecules per mL [MTM/mL] of plasma. Correlations between MTM/mL at diagnosis and ctDNA dynamics during NAT, residual cancer burden (RCB), and distant recurrence-free survival (DRFS) were examined across all subtypes.

**Results:** In all subtypes, high ctDNA concentration at diagnosis was associated with worse DRFS, whereas low ctDNA concentration or ctDNA-negative status was associated with improved DRFS, even with high tumor burden after NAT (RCB-II/RCB-III). We also found that patients with high ctDNA concentration, regardless of subtype, were less likely to experience early ctDNA clearance; however, those who did had a significantly higher likelihood of achieving a favorable response (RCB-0/RCB-I) than those with late or no ctDNA clearance. Furthermore, across all subtypes, patients with early ctDNA clearance, including those with substantial residual cancer (RCB-II/RCB-III) after NAT, had improved DRFS, irrespective of the ctDNA concentration at diagnosis.

**Conclusions:** Across all subtypes, pathologic response and ctDNA clearance reduce the risk of distant recurrence associated with high ctDNA concentration at diagnosis. ctDNA concentration at diagnosis and ctDNA clearance dynamics during NAT may facilitate the prediction of treatment response and further stratify the risk of metastatic recurrence in non-responders.

Trial Registration: NCT01042379

## INTRODUCTION

An accurate estimation of the risk of distant metastatic recurrence could facilitate the management of early-stage breast cancer ^1^. Risk assessment using prognostic biomarkers could inform treatment selection for personalized medicine. For example, patients at high risk of metastatic recurrence could be offered more aggressive treatment (escalation) to prevent or slow down the spread of cancer. In contrast, those with a low risk of relapse could forgo additional treatment (de-escalation), especially if their response to treatment was favorable, to avoid the toxicity of unnecessary therapies.

Measures of tumor burden (e.g., clinical T and N stages) and tumor characteristics (MammaPrint score, tumor grade, and subtype) evaluated during diagnostic workup have been used to estimate the risk of metastatic recurrence in early-stage breast cancers ^2, 3^. Circulating tumor DNA (ctDNA) is an emerging measure of tumor burden with strong prognostic value ^4^. Meta-analyses ^5, 6^, including previous studies from our group ^7, 8^, have unequivocally demonstrated that ctDNA positivity (as a binary variable) is associated with an increased risk of distant metastasis. However, most women with molecularly high-risk disease undergo neoadjuvant therapy (NAT), providing the opportunity to evaluate the more relevant assessment, which is the response to therapy. The significance of ctDNA concentration (as a continuous variable) remained underexplored in this context.

This study builds on our prior work showing that ctDNA status and clearance predict treatment response and recurrence risk in patients with high-risk early-stage breast cancer enrolled in the I-SPY2 trial ^7–9^. Unlike the published studies ^7–9^, which focused primarily on binary ctDNA status (positive/negative) and ctDNA dynamics (timing of clearance), the present study evaluates ctDNA concentration at diagnosis as a risk factor in the context of NAT response. Specifically, we investigated whether categorizing ctDNA-positive patients by ctDNA concentration could refine risk stratification and improve prediction of residual cancer burden (RCB). To account for heterogeneity in response rates and survival across breast cancer receptor subtypes, analyses were performed within hormone receptor (HR)-positive/HER2-negative, triple-negative (TN), and human epidermal growth factor receptor 2 (HER2)-positive breast cancers. We hypothesized that ctDNA concentration may provide additional information beyond the binary variable (i.e., ctDNA-positive vs. ctDNA-negative) for predicting treatment response and estimating the risk of metastatic recurrence.

## METHODS

### Patients, treatment, and samples

The study included 712 patients with MammaPrint ^10^ high-risk early-stage breast cancer enrolled in the neoadjuvant I-SPY2 trial (NCT01042379), a subset of the previously described 723-patient cohort ^9^ (**Table S1**). Patients received investigational agents combined with paclitaxel, followed by doxorubicin/cyclophosphamide (**Table S2, Supplementary Methods**). The Institutional Review Boards of all participating institutions approved the clinical research protocol (see **Supplementary Material**). All patients provided written informed consent to allow research on their biospecimen samples.

### ctDNA analysis

A personalized tumor-informed assay (Signatera^TM^, Natera, Inc.) ^7, 8^ was used to detect ctDNA in plasma samples collected at diagnosis (pretreatment, T0), at weeks 3 (T1) and 12 (T2) after treatment initiation, and post-NAT before surgery (T3). ctDNA concentration was reported as mean tumor molecules per mL [MTM/mL] of plasma. Patients with complete ctDNA data (**Table S3**) were grouped by ctDNA dynamics (**Supplementary Methods**). For this analysis, tertiles were established based on ctDNA concentration [low (tertile 1); intermediate (tertile 2); high (tertile 3)]; however, this categorization was not intended to develop guidelines for clinical or other research applications.

### Statistical analysis

We examined the correlation between ctDNA concentration at diagnosis and ctDNA dynamics, as well as distant recurrence-free survival (DRFS). Kaplan-Meier plots were constructed to visualize survival curves, which were then compared using the log-rank test. Univariable and multivariable Cox regression analyses were performed to estimate hazard ratios and their 95% confidence intervals (CI). P values were calculated using the Wald test.

We also assessed the association between ctDNA dynamics and residual cancer burden (RCB) ^11^ after NAT. Logistic regression was used to estimate odds ratios (OR) and 95% CIs. P values were calculated using the likelihood ratio (LR) test.

Statistical tests and data visualization were conducted using R packages ^12^. A P value <0.05 was considered statistically significant.

## RESULTS

### Patient characteristics at the time of diagnosis

Among 712 newly diagnosed cases of MammaPrint high-risk early-stage breast cancer (**Fig. S1**), 293 (41.2%) were HR-positive/HER2-negative, 236 (33.1%) were TN, and 183 (25.7%) were HER2-positive (**Table S1**). A total of 210 (30%) patients had clinical T3/T4 tumors; 392 (56.7%) were node-positive; 368 (71.2%) had grade 3 disease; and 393 (55.2%) had MammaPrint high 2 tumors. The patients received NAT in the I-SPY2 trial (**Table S2**), which evaluated the efficacy of various investigational agents in combination with chemotherapy.

### ctDNA concentration at diagnosis is associated with aggressive breast tumor characteristics

At baseline, 81.3% of patients (579/712) were ctDNA-positive. For the purposes of this analysis, the ctDNA-positive patients were divided into tertiles (n=193 per group) with increasing MTM/mL: low (tertile 1), intermediate (tertile 2), and high (tertile 3) (**Fig. S2**). We then examined whether ctDNA concentration at diagnosis, treated as a continuous or categorical variable (i.e., ctDNA concentration tertiles), was associated with clinicopathologic characteristics. Patients with TN disease, larger tumors (T3/T4), node-positive disease, higher grade (grade 3), and MammaPrint scores (high 2) were significantly enriched for patients in the high (tertile 3) ctDNA concentration group (all Chi-squared P <0.001, **Fig. 1A**). We found significantly higher median MTM/mL in the TN subtype, in patients with larger tumors (T3/T4), node-positive disease, higher grade (grade 3), and MammaPrint scores (high 2) (**Fig. 1B**). The same analyses conducted across subtypes yielded similar results (**Fig. S3, Fig. S4**).

**Figure 1.**
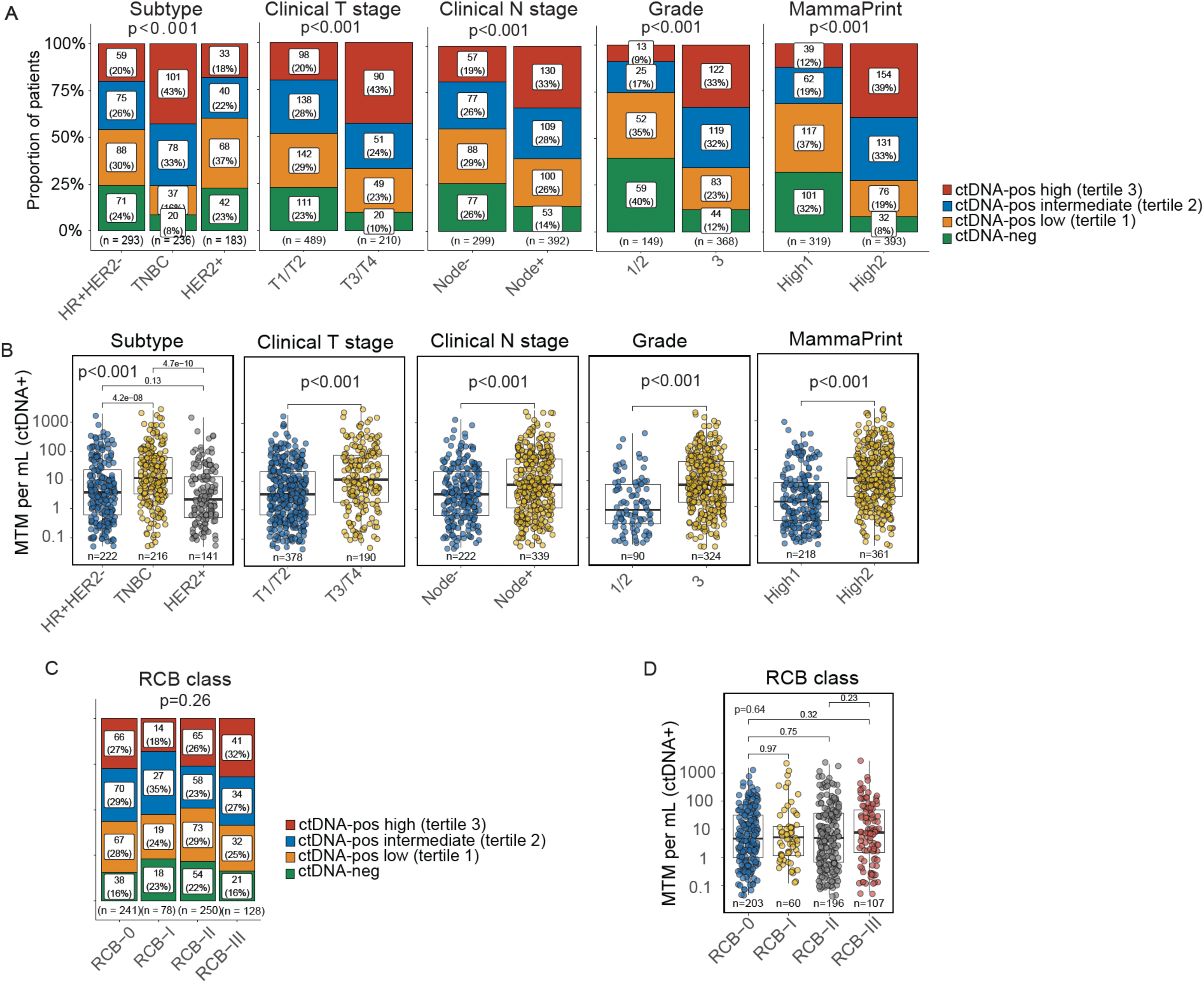
Association between ctDNA concentration at diagnosis and pretreatment tumor characteristics and treatment response. **A.** Bar plots show the proportions of patients grouped by ctDNA concentration at diagnosis: ctDNA-negative and ctDNA-positive: low (tertile 1), intermediate (tertile 2), and high (tertile 3). The clinicopathologic characteristics examined include subtype (HR+HER2- – hormone receptor- positive/HER2-negative vs. TNBC – triple-negative breast cancer vs. HER2+ – HER2-positive), clinical T stage (T1/T2 vs. T3/T4), clinical N stage (node-negative vs. node-positive), grade (1/2 vs. 3), and MammaPrint score (high 1 vs. high 2). P values were calculated using the Chi-squared test. The same analyses performed on each HR/HER2 subtype are shown in **Figure S3**. **B.** Box-and-whisker and dot plots show the distribution of ctDNA concentration at diagnosis, expressed as mean tumor molecules per milliliter (MTM/mL), by clinicopathologic characteristics. Each dot represents a patient’s ctDNA concentration at diagnosis. The same analyses performed on each HR/HER2 subtype are shown in **Figure S4**. **C.** Bar plot shows the proportions of patients grouped by ctDNA concentration at diagnosis: ctDNA-negative and ctDNA-positive: low (tertile 1), intermediate (tertile 2), and high (tertile 3) across the residual cancer burden (RCB) classes. P values were calculated using the Chi-squared test. The same analyses performed on each HR/HER2 subtype are shown in **Figure S3**. **D.** Box-and-whisker and dot plot show the distribution of ctDNA concentration at diagnosis, expressed as mean tumor molecules per milliliter (MTM/mL), by RCB class. Each dot represents a patient’s ctDNA concentration at diagnosis. The same analyses performed on each HR/HER2 subtype are shown in **Figure S4**. Within each box plot, the middle horizontal line indicates the median; the boxes span the 25th to the 75th percentile of each group’s distribution of values, and vertical lines from the boxes (whiskers) typically extend to the most extreme data points within 1.5 times the interquartile range (IQR) from the respective quartiles. P values were calculated using the Kruskal-Wallis test (>2 groups) or the Wilcoxon rank-sum test (2 groups).

### ctDNA concentration at diagnosis is not associated with treatment response, except in TN

Next, we investigated whether ctDNA concentration at diagnosis was associated with treatment response, as measured by the RCB method, at the time of surgery. Analysis across all patients revealed no significant association between RCB class and ctDNA concentration at diagnosis, either as a categorical variable (Chi-squared p=0.26, **Fig. 1C**) or as a continuous variable (Kruskal-Wallis p=0.64, **Fig. 1D**). However, analysis across subtypes revealed significant associations in the TN group, where patients with higher ctDNA concentration at baseline, either as a categorical (Chi-squared p=0.003, **Fig. S3**) or continuous (Wilcoxon p=0.0066, **Fig. S4**) variable were more likely to have extensive residual disease (RCB-III) than those with lower initial concentrations.

### Grouping patients by ctDNA concentration at diagnosis refines risk stratification in the context of RCB

Of the 712 patients, 133 (18.7%) experienced a DRFS event during a median follow-up of 4.7 years. As expected, RCB class was strongly associated with DRFS, where patients with RCB-0/RCB-I had excellent outcomes (**Fig. 2A**). As previously shown ^7, 9^, patients who tested ctDNA-positive at diagnosis (**Fig. S5**) had significantly worse DRFS than those who tested ctDNA-negative. Based on the observation that further stratification of ctDNA-positive patients into tertiles provided additional prognostic information (**Fig. 2B**), we investigated whether ctDNA concentration at diagnosis can further stratify risk of metastatic recurrence in the context of RCB.

**Figure 2.**
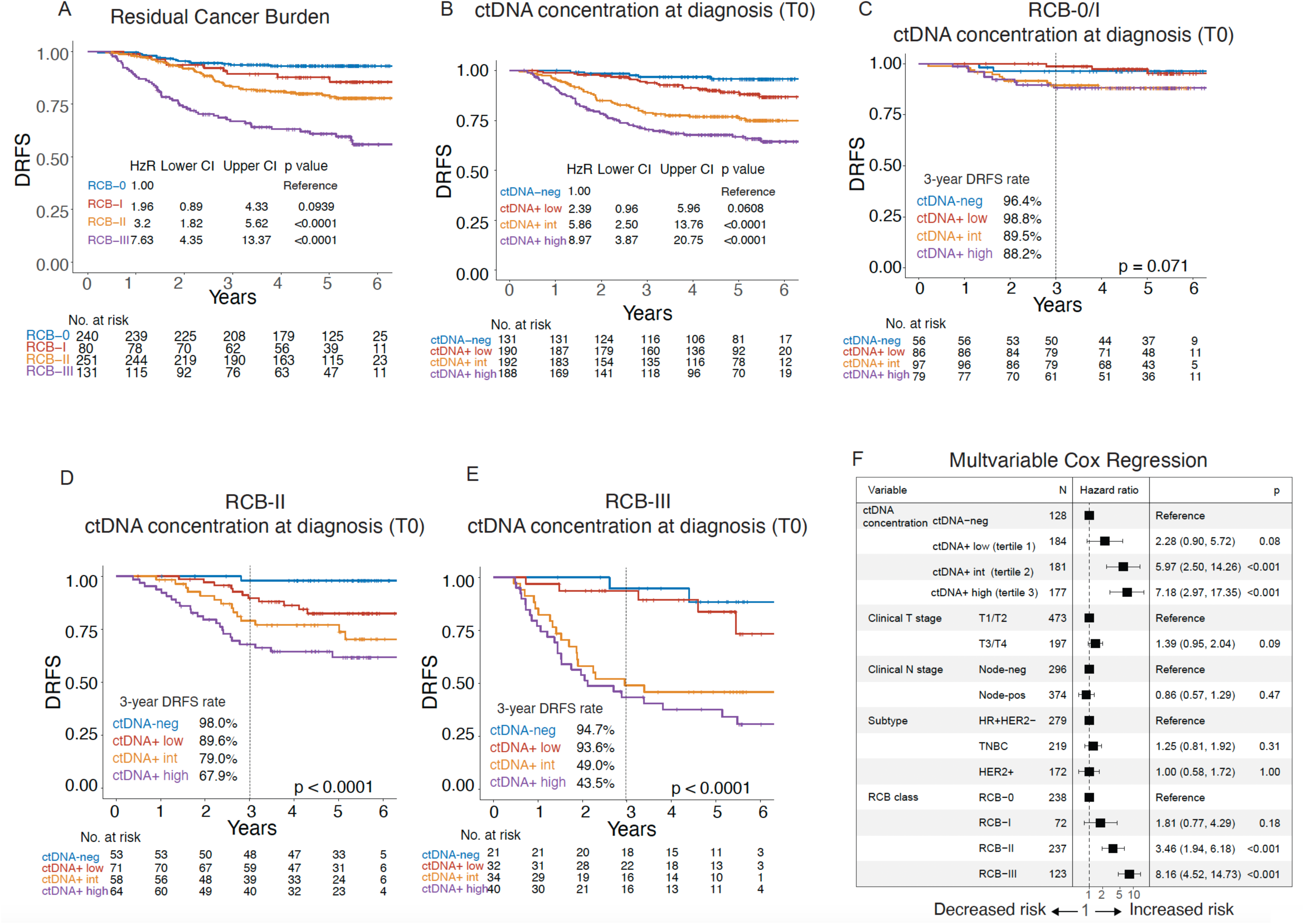
Impact of ctDNA concentration at diagnosis on distant metastatic recurrence in the context of treatment response. Kaplan-Meier plots compare distant recurrence-free survival (DRFS) of patients stratified according to. **A.** residual cancer burden (RCB): RCB-0 (pathologic complete response), RCB-I (limited), RCB-II (moderate), RCB-III (extensive) **B.** ctDNA concentration at diagnosis: ctDNA-negative and ctDNA positive: low (tertile 1), intermediate (tertile 2), and high (tertile 3). The hazard ratios and 95% confidence intervals (CI) from univariable Cox regression analyses are shown at the bottom of each survival plot. P values were calculated using the Wald test. **C-E**. Patients in each ctDNA concentration group were stratified by residual cancer burden (RCB) class: **C.** RCB-0/RCB-I; **D.** RCB-II; or **E.** RCB-III. The 3-year DRFS rates are shown in the bottom-left quadrant of each plot. P values were calculated using the log-rank test. The same analyses shown in **B.**, **C.**, and combined RCB-II/RCB-III were performed across HR/HER2 subtypes (**Figure S6**). **F.** Forest plot showing hazard ratios and 95% confidence intervals from multivariable Cox regression analysis, adjusted for clinicopathologic variables significantly associated with DRFS in univariable analyses (**Figure S5**). P values were calculated using the Wald test.

Patients with RCB-0/RCB-I had favorable survival regardless of ctDNA concentration group at diagnosis (3-year DRFS rate >88%, **Fig. 2C**). However, in patients with residual disease at surgery [RCB-II (**Fig. 2D**) or RCB-III (**Fig. 2E**)], we found that applying the tertiles further refined risk stratification such that those in the intermediate (tertile 2) and high (tertile 3) groups had significantly worse DRFS than their ctDNA-negative counterparts (all log-rank P <0.001). Conversely, those in the ctDNA-negative or low (tertile 1) group had favorable survival (3-year DRFS rate >89%) despite substantial disease remaining after NAT (RCB-II or RCB-III). These findings were similar across receptor subtypes (**Fig. S6**). In a Cox multivariate model that included baseline clinical variables, both pretreatment ctDNA concentration and RCB class remained significantly and independently associated with DRFS (**Fig. 2F, Fig. S5**).

### Early ctDNA clearance in patients with high ctDNA concentration at diagnosis is associated with good response

We first examined whether ctDNA clearance dynamics depend on the initial ctDNA concentration at diagnosis across subtypes. The ctDNA clearance patterns as a function of initial concentration grouping are shown for the overall cohort (**Fig. 3A**) and by subtype (**Fig. 3B**). Logistic regression analysis showed a significantly lower likelihood of early ctDNA clearance at week 3 (T1) in patients in the high (tertile 3) ctDNA concentration group compared with those in the low (tertile 1) ctDNA concentration group (OR 0.12, 95% CI 0.04-0.29, LR P <0.001, **Fig. 3C**). The signal was strongest in the HR-positive/HER2-negative subtype (OR 0.15, 95% CI 0.03-0.55, LR p=0.007, **Fig. 3D**).

**Figure 3.**
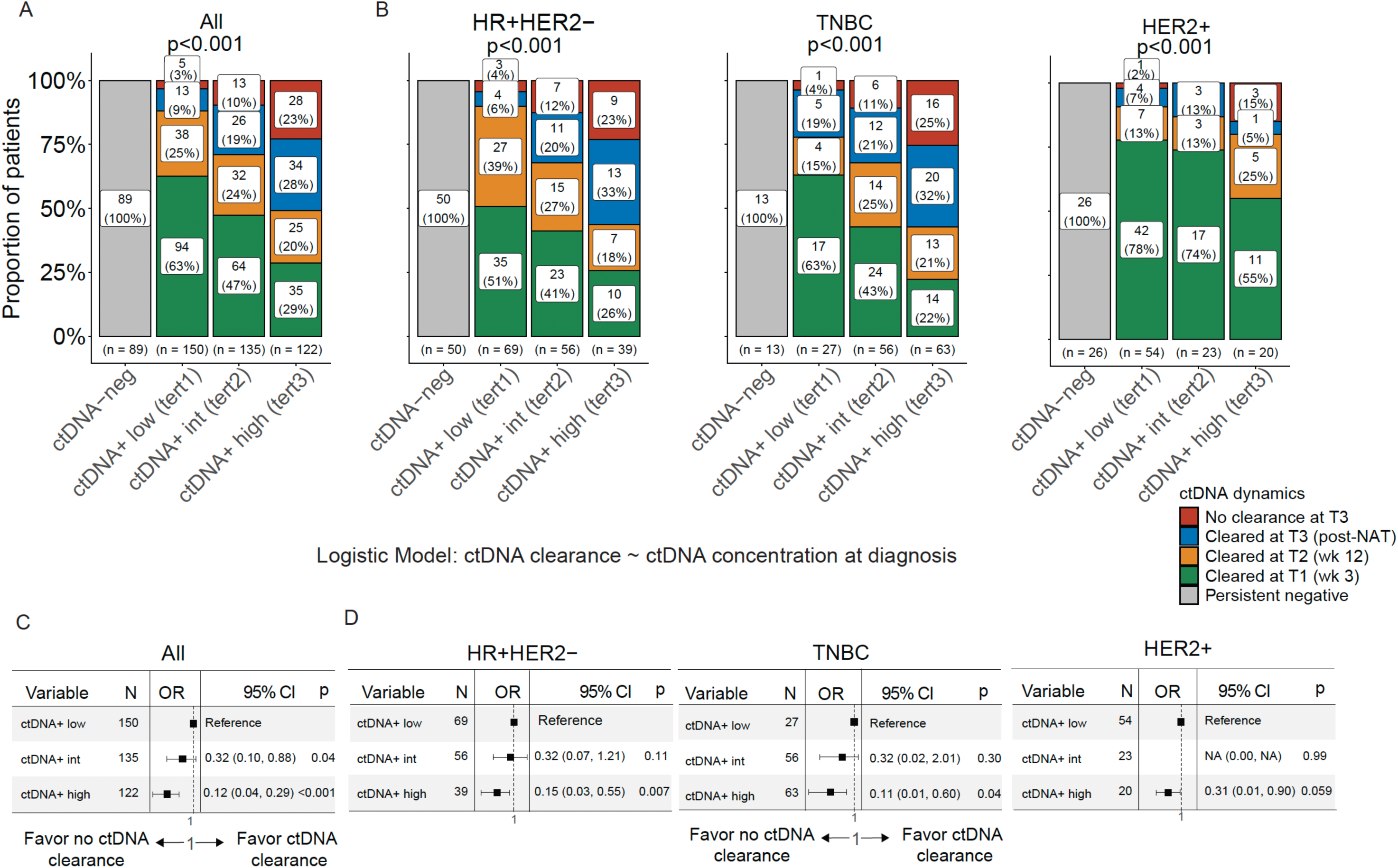
Relationship between ctDNA concentration at diagnosis and likelihood of ctDNA clearance. **A.** Bar plots show the proportions of patients by ctDNA dynamics: persistent negative, cleared at week 3 (T1) or week 12 (T2), or post-neoadjuvant therapy (T3), and no clearance at T3 stratified by ctDNA concentration at diagnosis: ctDNA-negative and ctDNA positive: low (tertile 1), intermediate (tertile 2), and high (tertile 3). **B.** The same analysis was performed across subtypes: hormone receptor-positive/HER2-negative (HR+HER2-), triple-negative breast cancer (TNBC), and HER2-positive (HER2+). **C.** Logistic regression was performed in all patients to estimate odds ratios (OR) and 95% confidence intervals (CI) for ctDNA clearance (cleared at T1, T2, or T3) vs. no clearance at T3. P values were calculated using the likelihood ratio test. **D.** The same analysis was performed across HR/HER2 subtypes.

Next, we investigated whether ctDNA dynamics within ctDNA concentration groups were associated with treatment response. The RCB distributions as a function of clearance dynamics within baseline ctDNA concentration groups are shown in **Fig. 4A**. For the full cohort, logistic regression analysis revealed that ctDNA clearance was significantly associated with an increased likelihood of RCB-0/RCB-I, regardless of pretreatment ctDNA concentration group (all LR P <0.05, **Fig. 4B**). Across all subtypes, early ctDNA clearance was associated with better response (lower RCB class) in patients with intermediate or high ctDNA concentration at diagnosis (**Fig. S7**).

**Figure 4.**
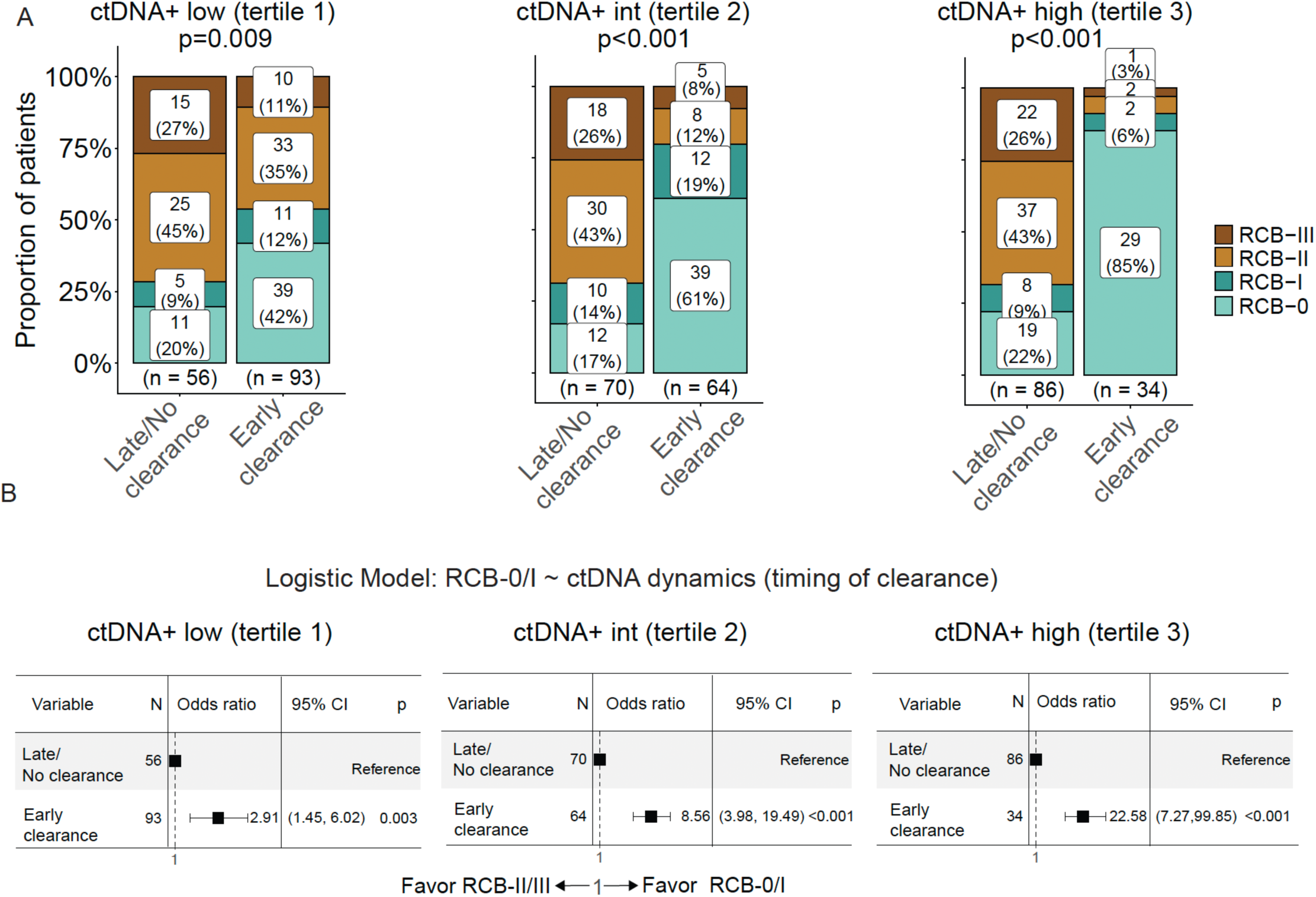
Early ctDNA clearance enriches for good responders. Patients who were ctDNA-positive at diagnosis were grouped by ctDNA concentration at diagnosis: low (tertile 1), intermediate (tertile 2), and high (tertile 3). **A.** Bar plots show the proportions of patients and their residual cancer burden (RCB) class, grouped by ctDNA dynamics: Early clearance vs. Late or No clearance. ‘Early clearance’ included ctDNA-positive patients with ctDNA clearance at week 3 after treatment initiation (T1). ‘Late or no clearance’ included those with ctDNA clearance at week 12 (T2), or post-neoadjuvant therapy (T3), or no clearance at T3. P values were calculated using the Chi-squared test. The same analysis was performed across HR/HER2 subtypes (**Figure S7**). **B.** Logistic regression was performed to estimate odds ratios and 95% confidence intervals for achieving RCB- 0/RCB-I vs. RCB-II/RCB-III based on ctDNA dynamics in patients with ‘Early clearance’ vs. ‘Late or no clearance’.

### Early ctDNA clearance mitigates the risk of distant metastatic recurrence conferred by high initial ctDNA concentrations in non-responders

Our prior studies showed that ctDNA dynamics are associated with the risk of metastatic recurrence among poor responders ^7, 9^. Here, we examined whether ctDNA dynamics are prognostic for DRFS within each ctDNA concentration group in patients with moderate (RCB-II) or extensive RCB (RCB-III) after NAT. Patients with early ctDNA clearance [at weeks 3 (T1) or 12 (T2)] had significantly higher DRFS rates compared to those without ctDNA clearance post-NAT (T3) regardless of the ctDNA concentration group (all log-rank P <0.05, **Fig. 5A**). This correlation was observed in the full cohort and across subtypes (all log-rank P <0.05, **Fig. S8**, **Fig. S9, Table S3**). Early ctDNA clearance was significantly associated with a lower risk of metastatic recurrence across all tertiles (**Fig. 5B**). Thus, among poor responders, patients who clear their ctDNA by weeks 3 or 12 after treatment start have improved DRFS, irrespective of the initial ctDNA concentration at diagnosis.

**Figure 5.**
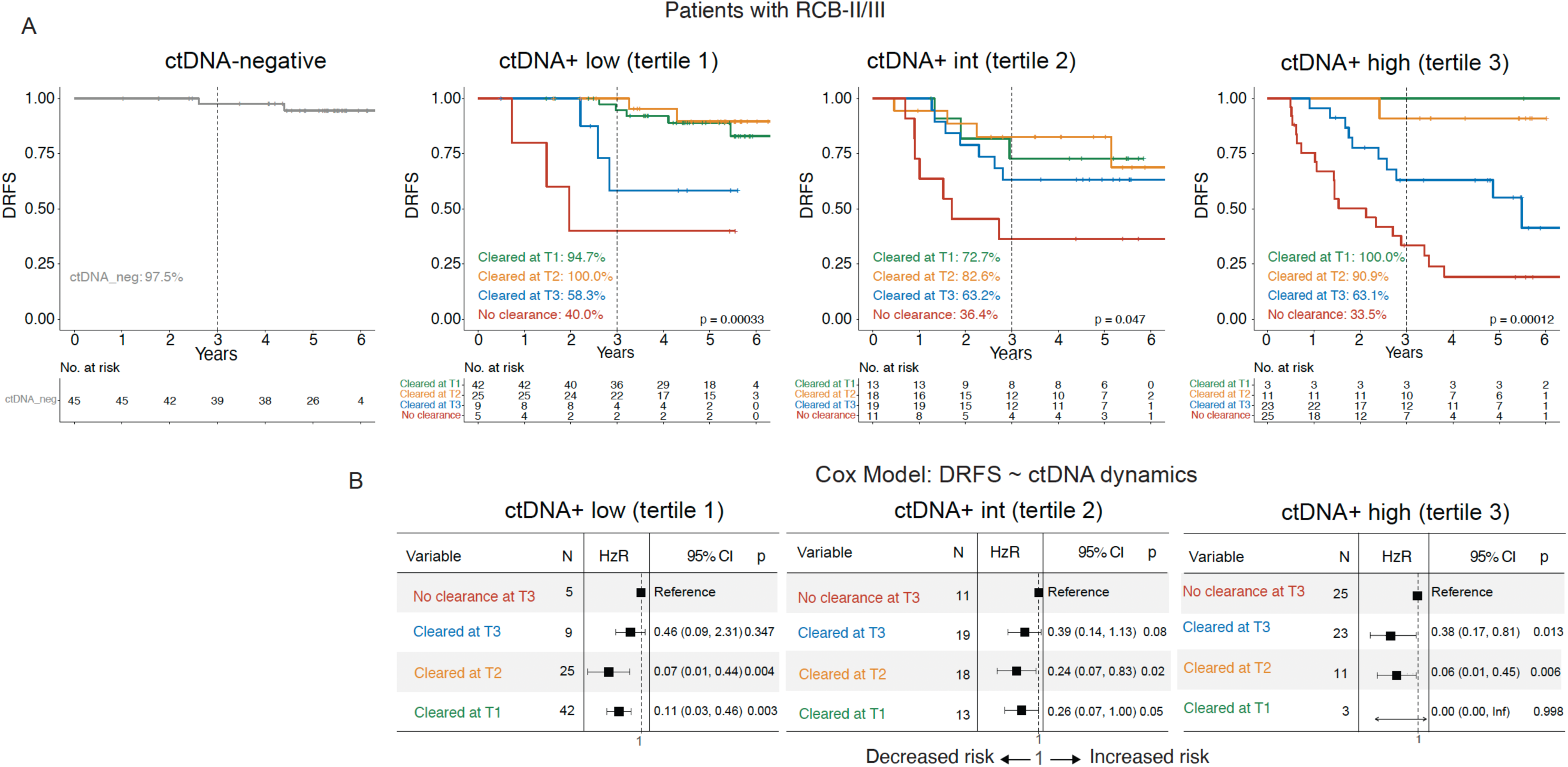
Correlation between ctDNA clearance and the risk of metastatic recurrence in patients with moderate or extensive residual cancer burden (RCB-II or RCB-III) after neoadjuvant therapy (NAT) stratified by ctDNA concentration at diagnosis. **A.** Kaplan-Meier plots compare distant recurrence-free survival (DRFS) across 4 patient groups by ctDNA concentration at diagnosis (columns): ctDNA-negative and ctDNA positive: low (tertile 1), intermediate (tertile 2), and high (tertile 3). Each ctDNA concentration group was stratified by ctDNA dynamics: persistent negative, cleared at week 3 (T1) or week 12 (T2) after treatment initiation, cleared at post-NAT before surgery (T3), and no clearance at T3. The 3-year DRFS rates, according to ctDNA dynamics, are shown in the lower-left quadrant. The P value was calculated using the log-rank test. To exclude the confounding effects of favorable DRFS in patients with RCB-0 (pathologic complete response) or RCB-I (limited) after NAT, the analysis was performed only in patients with RCB-II (moderate) or RCB-III (extensive). **B.** Cox regression analysis was performed to estimate hazard ratios and 95% confidence intervals (CI) for ctDNA-positive patients grouped by ctDNA dynamics: cleared at T1, T2, or T3, and no clearance at T3. The no clearance group served as the reference.

## DISCUSSION

This study examined the impact of ctDNA concentration at diagnosis and ctDNA dynamics during treatment on NAT response and the risk of distant metastatic recurrence. As expected, we found a significant association between high ctDNA concentration at diagnosis and clinicopathologic characteristics indicative of aggressive breast cancer. Examining the association of ctDNA concentration at diagnosis with NAT response, we found that in the full cohort, ctDNA concentration at diagnosis was not associated with treatment response (RCB class). However, there was an association between high ctDNA concentration at diagnosis and poor response (RCB-II/RCB-III) in the TN subset. Consistent with previous work ^7–9^, we also found that early ctDNA clearance was associated with a significantly higher likelihood of achieving a good response to NAT (RCB- 0/RCB-I), including in patients with high ctDNA concentration at diagnosis.

Consistent with results from numerous clinical studies, lower RCB (RCB-0/RCB-I) and ctDNA-negativity were significantly associated with improved DRFS. Here, we investigated whether ctDNA concentration at diagnosis can further stratify risk of recurrence in the context of RCB and ctDNA clearance dynamics. We found that, among patients with RCB-II or RCB-III, ctDNA concentration at diagnosis added value by further stratifying patients into prognostic groups and identifying those with low ctDNA concentration at diagnosis as having favorable DRFS, despite having substantial residual cancer after NAT. Moreover, non-responders (RCB-II/RCB-III) with early ctDNA clearance had improved DRFS, even among those with high ctDNA concentration at diagnosis.

The increased risk of distant recurrence associated with high ctDNA concentration at diagnosis was mitigated by two factors: low disease burden after NAT (RCB-0/RCB-I) and early ctDNA clearance, both of which may be considered as indicators of a good response to NAT. Patients with an excellent tumor response to NAT (RCB-0/RCB-I) have improved outcomes, as do those who clear ctDNA early, regardless of initial ctDNA concentration. This aligns with our previous findings demonstrating improved DRFS in patients with early ctDNA clearance ^7–9^, as well as our earlier conclusion that a high disease burden can be mitigated by an excellent response to NAT ^13^.

Adjusting treatments based on early ctDNA clearance, especially in patients with high ctDNA concentration at diagnosis, could improve patient outcomes. These findings support prospective evaluation of ctDNA-guided escalation and de-escalation strategies. For example, patients who do not clear by week 3 could be offered more aggressive treatment (escalation) to facilitate subsequent clearance of ctDNA. Upon ctDNA clearance, patients may receive surgery early, forgoing additional treatments (de-escalation) that could expose them to the toxicity of unnecessary therapies. In I-SPY2, we have ongoing studies looking at the role of ctDNA clearance in improving predictions based on imaging and on-treatment biopsy. ctDNA dynamics may be helpful, particularly in identifying those who could benefit from switching earlier to different therapies.

Recent studies from our group ^7–9, 14^ and others ^15–24^ have demonstrated the clinical significance of ctDNA in patients with breast cancer receiving NAT. These studies, including ours, have used binary test results (ctDNA-positive vs. ctDNA-negative) rather than ctDNA concentration as a prognostic and predictive biomarker. Another unit of measurement for ctDNA quantification, in addition to MTM/mL, is variant allele frequency (VAF), defined as the proportion of mutant DNA sequences relative to the total number of all sequences. A recent study in patients with TN disease showed that a baseline VAF ≥1.1% was associated with poor DRFS ^21^. Here, we used MTM/mL instead of VAF because it is normalized to the cell-free DNA yield (ng) extracted from plasma, which contains both ctDNA and DNA from dying hematopoietic cells ^25^. Cell-free DNA concentration in blood has been shown to have prognostic value ^26^, although its significance is less than that of ctDNA ^14^. We hypothesized that combining ctDNA levels (mutant DNA copies by VAF) with the cfDNA input amount (ng) to estimate ctDNA concentration (MTM/mL) would allow us to capture prognostic information from both measurements ^21^.

The study had several limitations. It included only patients at high risk (Stage 2 and 3, as well as MammaPrint high) for metastatic recurrence, and the findings may not be generalizable to lower-risk populations. While the full cohort was relatively large (n=712), further stratification yielded smaller subsets for analysis that could lead to chance findings.

In summary, we found that even in patients with high ctDNA concentration at diagnosis, early ctDNA clearance during NAT and/or an excellent response (RCB-0/RCB-I) were associated with excellent outcomes. However, in the absence of early ctDNA clearance or an excellent response, higher ctDNA concentrations are associated with a higher risk of metastatic recurrence, particularly in non-responding patients (RCB-II/RCB-III).

Ongoing analyses in I-SPY2 include modeling combinations of clinicopathologic variables, such as tumor response assessed by pathology and imaging ^27^, to optimize the prediction of RCB and DRFS. We are expanding the study to significantly increase the cohort size. Additionally, the new I-SPY2.2 trial design has implemented prospective serial ctDNA testing, which will be used in the future to guide treatment alongside imaging and pathology. Data from these efforts will be used to confirm the findings of this study, potentially enhancing our understanding of the clinical utility of ctDNA concentration at diagnosis and of ctDNA dynamics in the context of response, to guide treatment selection and improve outcomes for patients with high-risk early-stage breast cancer receiving NAT.

## Data Availability

De-identified subject-level data are made available to academic and not-for-profit investigators for projects approved by the I-SPY Data Access and Publications Committee. Details of the application and review process are available at: https://www.quantumleaphealth.org/for-investigators/clinicians-proposal-submissions/. Data is made available to approved applicants within 1 to 2 months of application.

## ACKNOWLEDGMENTS

This research was supported by the National Cancer Institute (NCI) of the National Institutes of Health (NIH) under award R01CA255442. Additional funding was provided by NCI NIH award number P01CA210961. The authors wish to acknowledge the generous support of the study sponsors, Quantum Leap Healthcare Collaborative (QLHC, 2013 to present), and the Foundation for the National Institutes of Health (2010 to 2012). The authors sincerely appreciate the ongoing support for the I-SPY2 Trial from the Safeway Foundation, the William K. Bowes, Jr. Foundation, Give Breast Cancer the Boot, and the Breast Cancer Research Foundation. We thank Minetta Liu, Angel Rodriguez, and Philip Miller (Natera, Inc) for the review of the manuscript. Sincere thanks to all the patients who have volunteered to participate in I-SPY2. This study is in collaboration with Merck Sharp & Dohme LLC, a subsidiary of Merck & Co., Inc., Rahway, NJ, USA.

