## Supplementary Information for "Circulating tumor DNA concentration at diagnosis is a modifiable prognostic factor for distant metastatic recurrence in patients with high-risk breast cancer receiving neoadjuvant therapy"

**Table of Contents**

| | Page |
|---|---|
| Authors | 2 |
| Supplementary Methods | 3 |
| Figure S1. Consort flow chart. | 5 |
| Figure S2. ctDNA concentration at diagnosis in patients with high-risk early-stage breast cancer. | 6 |
| Figure S3. Grouping by ctDNA concentration at diagnosis and its association with tumor characteristics and treatment response across subtypes | 8 |
| Figure S4. ctDNA concentration at diagnosis and its association with tumor characteristics and treatment response. | 10 |
| Figure S5. Distant recurrence-free survival in patients grouped according to tumor characteristics at diagnosis. | 12 |
| Figure S6. ctDNA concentration at diagnosis is a prognostic factor for distant metastatic recurrence. | 14 |
| Figure S7. Patients across subtypes with intermediate or high ctDNA concentration at diagnosis who clear ctDNA are significantly enriched for good responders | 16 |
| Figure S8. Correlation between ctDNA clearance and risk of metastatic recurrence in all patients (complete ctDNA data) stratified by ctDNA concentration at diagnosis. | 17 |
| Figure S9. Correlation between ctDNA clearance and risk of metastatic recurrence across subtypes stratified by ctDNA concentration at diagnosis. | 19 |
| Table S1. Patient and tumor characteristics. | 21 |
| Table S2. Treatment received in the I-SPY2 trial. | 23 |
| Table S3. Patients with complete ctDNA data at all four time points. | 24 |
| References | 25 |

**Supplementary Methods**

**Patients, treatment, and samples.** Patients with MammaPrint high-risk early-stage breast cancer who received neoadjuvant therapy (NAT) in the I-SPY2 trial (NCT01042379) were included in the study (**Table S1**). MammaPrint is a prognostic assay that classifies breast cancers into high- and low-risk subgroups [1]. The high-risk group can be further subdivided into high-risk (high-1) and very high-risk (high-2).

Patients received 12 weekly cycles of paclitaxel alone or in combination with an investigational drug, followed by 4 3-week cycles of doxorubicin/cyclophosphamide (**Table S2**). Patients who have HER2-positive disease also received trastuzumab for the first 12 weeks.

**ctDNA analysis**. ctDNA testing was performed on blood samples collected at diagnosis (pretreatment, T0), at week 3 (T1), at week 12 (T2) after treatment initiation, and post-NAT before surgery (T3). Detection of ctDNA in plasma was performed using a personalized tumor-informed ctDNA assay (SignateraTM, Natera Inc.). The assay used multiplexed polymerase chain reaction (PCR) to amplify up to 16 tumor-specific single-nucleotide variants (SNVs) from cell-free DNA (cfDNA). The PCR amplicons were then subjected to deep sequencing to determine variant allele frequency (VAF). A plasma sample was considered ctDNA-positive if ≥2 of the 16 variants were detected. The number of tumor molecules for each mutation was calculated by dividing the cfDNA input (ng) by the mass equivalent to the haploid human genome (0.003 ng) and multiplying by the VAF. The mean tumor molecules per mL (MTM/mL) was calculated by dividing the total number of tumor molecules detected in a sample by 16.

For this analysis, we grouped ctDNA-positive patients at diagnosis (n=579, 81%) into tertiles based on MTM/mL distribution (n=193 per tertile). Thus, the whole cohort was categorized into 4 ctDNA concentration groups: ctDNA-negative (n=133), and ctDNA-positive tertiles 1 (low, n=193), 2 (intermediate, n=193), and 3 (high, n=193).

In subset analyses of ctDNA dynamics during NAT, patients with complete ctDNA data at all 4 time points (**Table S3**) were grouped by the timing of ctDNA clearance:

a. persistent negative (ctDNA-/-/-/-);
b. cleared at week 3 (T1, ctDNA+/-/-/-);
c. cleared at week 12 (T2, ctDNA+/+/-/-);

d. cleared post-NAT before surgery (T3, ctDNA+/+/+/- or ctDNA+/-/+/-); or

e. no clearance at T3 (ctDNA+/+/+/+, ctDNA+/-/+/+, or ctDNA+/+/-/+).

**Statistical analysis.** The survival endpoint was distant recurrence-free survival (DRFS), defined as the time from treatment consent to the first distant recurrence or death from any cause. The response endpoint was residual cancer burden (RCB), defined as the amount of invasive cancer in the breast and regional lymph nodes after NAT, assessed by pathology at the time of surgery [2].

To assess associations with clinicopathologic variables, we compared the proportions of ctDNA groups or the median MTM/mL across subtypes (HR-positive/HER2-negative, TN and HER2-positive), clinical T stage (T1/T2 and T3/T4), clinical N stage (node-negative vs. node-positive), grade (1/2 and 3), MammaPrint score (high 1 and high 2), and RCB class (RCB-0/pathologic complete response [pCR], RCB-I, RCB-II, and RCB-III, representing no invasive cancer in the breast and regional lymph nodes, limited, moderate, and extensive RCB following NAT, respectively) [2]. P values were calculated using the Chi-squared test, Wilcoxon rank-sum test, or Kruskal-Wallis test.

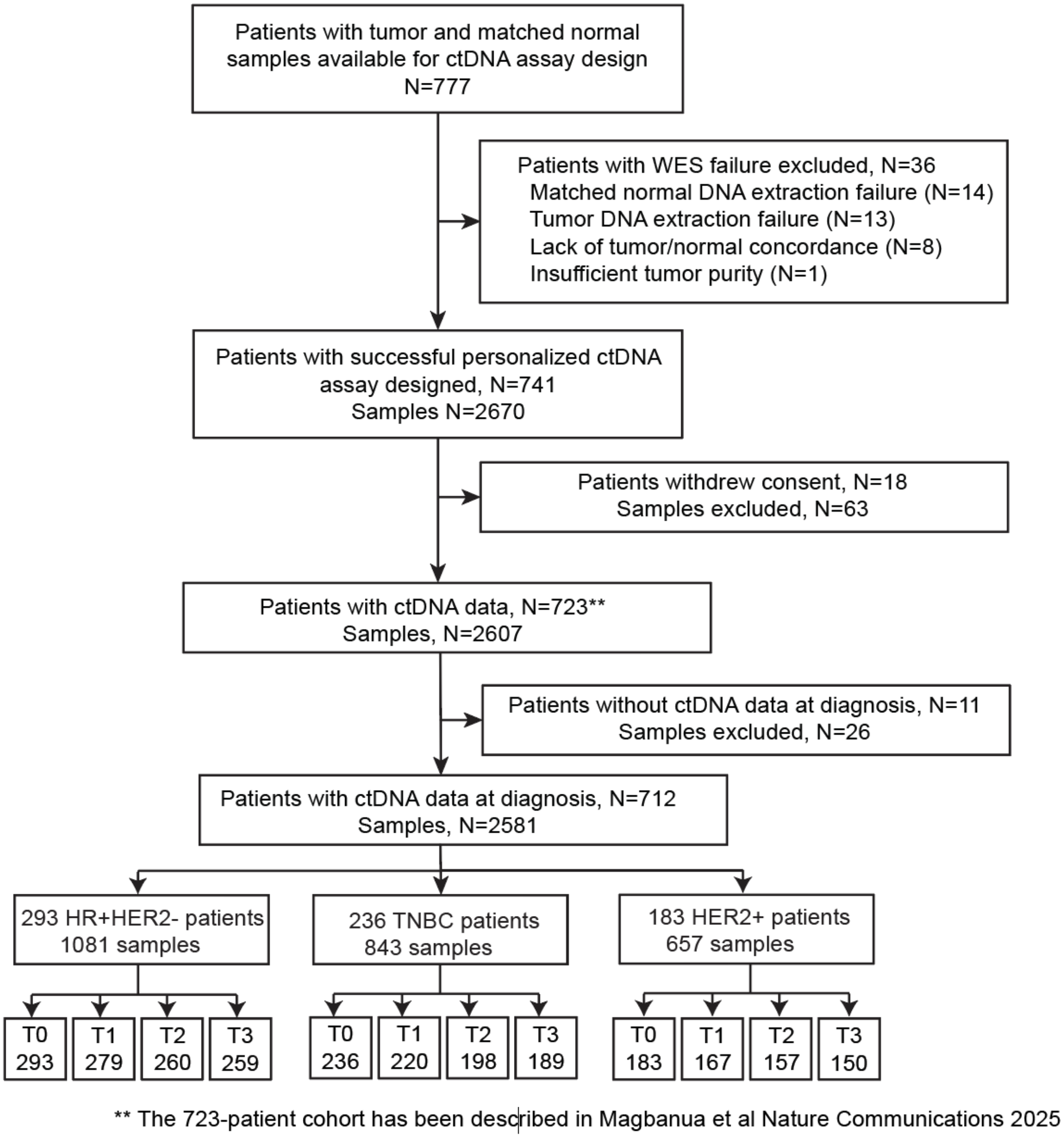

**Figure S1. Consort flow chart.** Diagram showing inclusion and exclusion of patients and samples in the study. This study includes 712 patients from the previously described 723-patient cohort . Abbreviations: HER2+ – HER2-positive, HR+HER2- – hormone receptor-positive/HER2-negative, T0 – at diagnosis (pretreatment), T1 – week 3 after treatment initiation, T2 – week 12 after treatment initiation, T3 – post-neoadjuvant therapy before surgery, TNBC – triple-negative breast cancer, WES – whole exome sequencing.

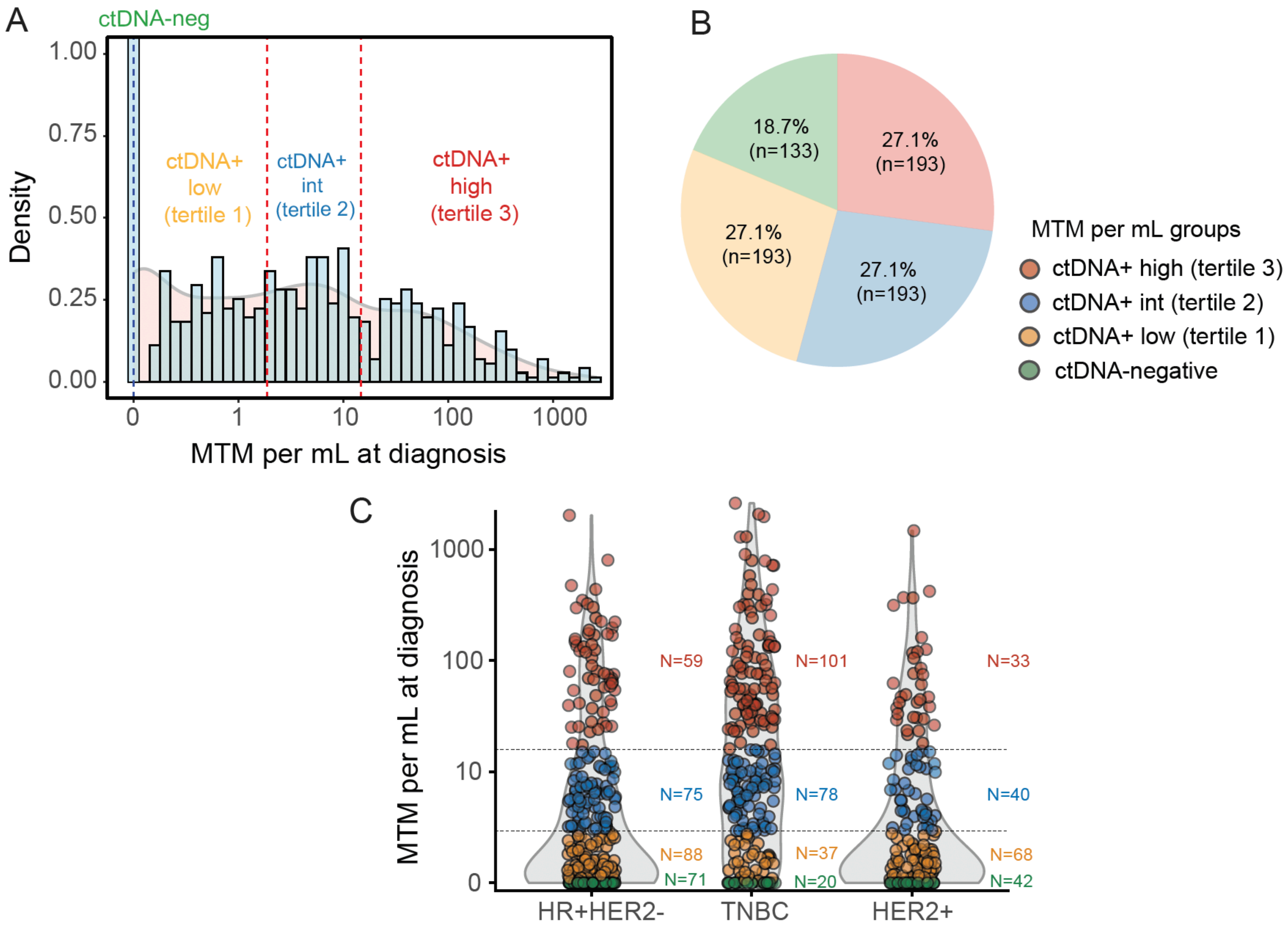
A
ctDNA-neg
Density
1.00
0.75
0.50
0.25
0.00
ctDNA+
low
(tertile 1)
ctDNA+
int
(tertile 2)
ctDNA+
high
(tertile 3)
0
1
10
100
1000
MTM per mL at diagnosis
B
18.7%
(n=133)
27.1%
(n=193)
27.1%
(n=193)
27.1%
(n=193)
MTM per mL groups
ctDNA+ high (tertile 3)
ctDNA+ int (tertile 2)
ctDNA+ low (tertile 1)
ctDNA-negative
C
MTM per mL at diagnosis
1000
100
10
0
N=59
N=75
N=88
N=71
N=101
N=78
N=37
N=20
N=33
N=40
N=68
N=42
HR+HER2-
TNBC
HER2+

**Figure S2. ctDNA concentration at diagnosis in patients with high-risk early-stage breast cancer.** ctDNA concentration in blood was measured using a personalized tumor-informed assay.

**A.** The distribution of mean tumor molecules per mL (MTM/mL) of plasma is shown as a histogram (blue). The density plot (pink) visualizes the distribution of continuous values (MTM/mL), with peaks showing where the values are concentrated. The total area under each distribution curve is equal to 1.

**B.** The ctDNA-positive patients were then divided equally into 3 groups based on increasing ctDNA concentration: low (tertile 1), intermediate (tertile 2), and high (tertile 3). A pie chart shows the percentages and numbers of patients in each ctDNA concentration group.

**C.** Violin and dot plots showing the distribution of MTM/mL by subtype based on hormone receptor (HR) and human epidermal growth factor 2 (HER2) status: HR+HER2- – HR-positive/HER2-negative, TNBC – triple-negative breast cancer, and HER2+ – HER2-positive. The number of patients (N) in each ctDNA concentration group is displayed to the right of each plot.

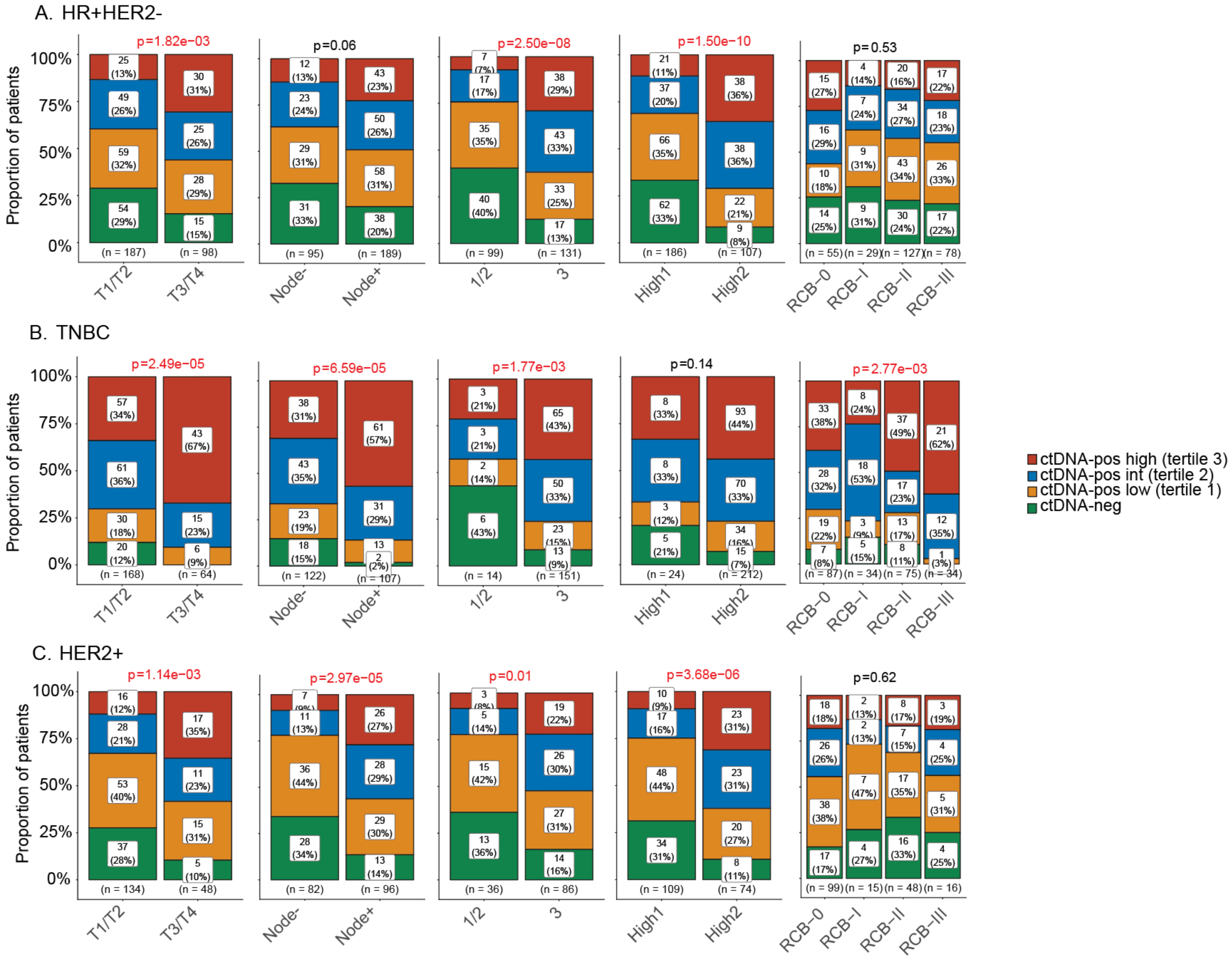

A. HR+HER2-
B. TNBC
C. HER2+
Proportion of patients
100%
75%
50%
25%
0%
p=1.82e−03
p=0.06
p=2.50e−08
p=1.50e−10
p=0.53
(n = 187) (n = 98)
(n = 95) (n = 189)
(n = 99) (n = 131)
(n = 186) (n = 107)
(n = 55) (n = 29) (n = 127) (n = 78)
p=2.49e−05
p=6.59e−05
p=1.77e−03
p=0.14
p=2.77e−03
(n = 168) (n = 64)
(n = 122) (n = 107)
(n = 14) (n = 151)
(n = 24) (n = 212)
(n = 87) (n = 34) (n = 75) (n = 34)
p=1.14e−03
p=2.97e−05
p=0.01
p=3.68e−06
p=0.62
(n = 134) (n = 48)
(n = 82) (n = 96)
(n = 36) (n = 86)
(n = 109) (n = 74)
(n = 99) (n = 15) (n = 48) (n = 16)
T1/T2
T3/T4
Node-
Node+
1/2
3
High1
High2
RCB−0
RCB−I
RCB−II
RCB−III
ctDNA-pos high (tertile 3)
ctDNA-pos int (tertile 2)
ctDNA-pos low (tertile 1)
ctDNA-neg

**Figure S3. Grouping by ctDNA concentration at diagnosis and its association with tumor characteristics and treatment response across subtypes.** Bar plots showing the proportions of patients grouped based on ctDNA concentration at diagnosis: ctDNA-negative and ctDNA positive: low (tertile 1), intermediate (tertile 2), and high (tertile 3). The clinicopathologic characteristics examined include subtype (HR+HER2- – hormone receptor-positive/HER2-negative vs. TNBC – triple-negative breast cancer, vs. HER2+ – HER2-positive), clinical T stage (T1/T2 vs. T3/T4), clinical N stage (node-negative vs. node-positive), grade (1/2 vs. 3), and MammaPrint score (high 1 vs. high 2), and residual cancer burden (RCB) class. The same analyses were performed on each subtype:

**A**. HR+HER2- – hormone receptor-positive/HER2-negative,

**B.** TNBC – triple-negative breast cancer, and

**C.** HER2+ – HER2-positive.

P values were calculated using the Chi-squared test. The red texts indicate statistically significant differences in the proportions of patients.

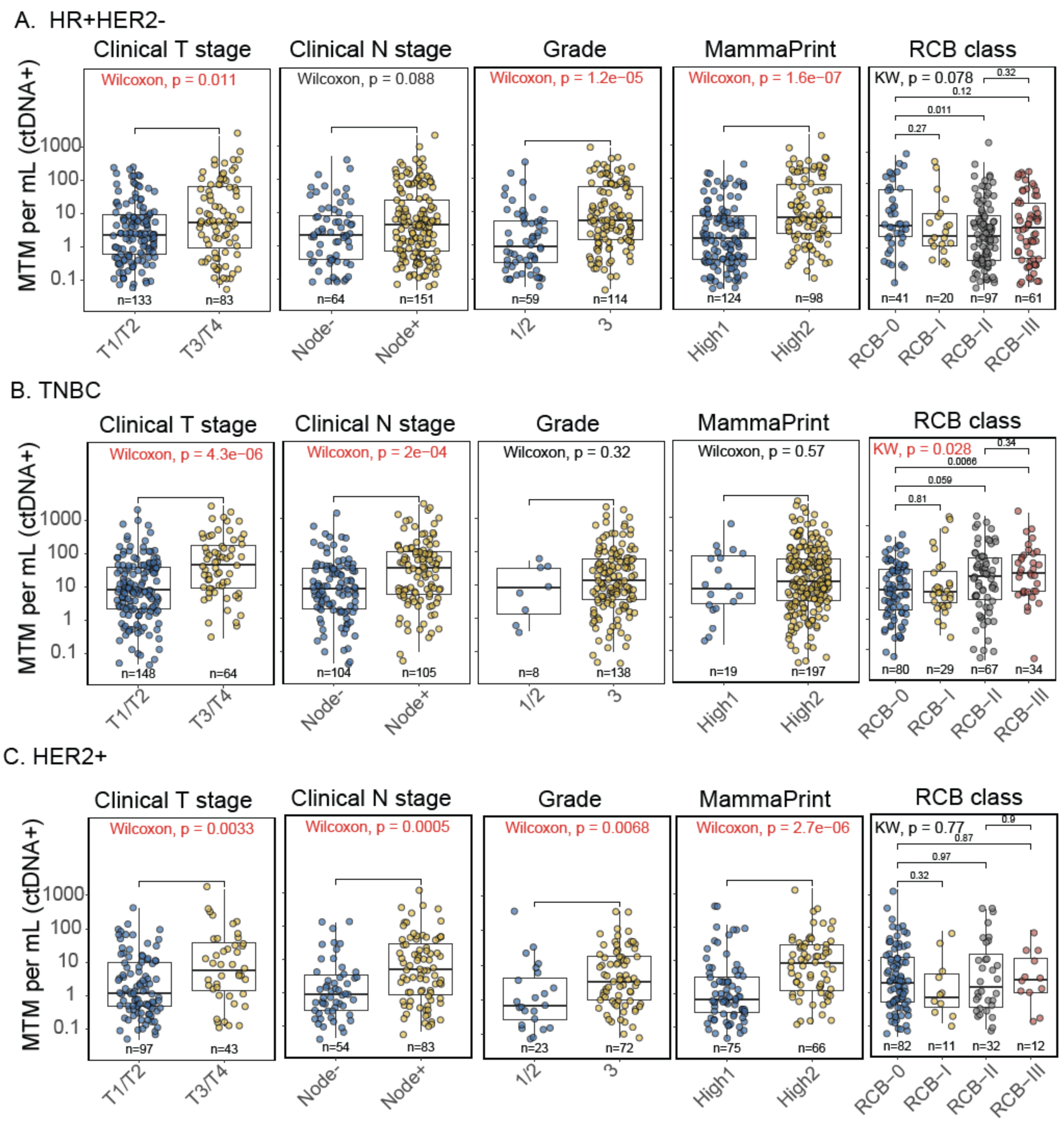
A. HR+HER2-
Clinical T stage
Clinical N stage
Grade
MammaPrint
RCB class
Wilcoxon, p = 0.011
Wilcoxon, p = 0.088
Wilcoxon, p = 1.2e−05
Wilcoxon, p = 1.6e−07
KW, p = 0.078
MTM per mL (ctDNA+)
n=133
n=83
n=64
n=151
n=59
n=114
n=124
n=98
n=41
n=20
n=97
n=61
T1/T2
T3/T4
Node-
Node+
1/2
3
High1
High2
RCB−0
RCB−I
RCB−II
RCB−III
B. TNBC
Wilcoxon, p = 4.3e−06
Wilcoxon, p = 2e−04
Wilcoxon, p = 0.32
Wilcoxon, p = 0.57
KW, p = 0.028
n=148
n=64
n=104
n=105
n=8
n=138
n=19
n=197
n=80
n=29
n=67
n=34
C. HER2+
Wilcoxon, p = 0.0033
Wilcoxon, p = 0.0005
Wilcoxon, p = 0.0068
Wilcoxon, p = 2.7e−06
KW, p = 0.77
n=97
n=43
n=54
n=83
n=23
n=72
n=75
n=66
n=82
n=11
n=32
n=12

**Figure S4. ctDNA concentration at diagnosis and its association with tumor characteristics and treatment response across subtypes.** Box-and-whisker and dot plots showing the distribution of ctDNA concentration at diagnosis expressed as mean tumor molecules per milliliter (MTM/mL) by clinicopathologic characteristics and RCB class. Each dot represents a patient's ctDNA concentration at diagnosis. Within each box, the middle horizontal line indicates the median; the boxes span from the 25th to the 75th percentile of each group's distribution of values, and vertical lines from the boxes (whiskers) typically extend to the most extreme data points within 1.5 times the interquartile range (IQR) from the respective quartiles. The same analyses performed on each subtype:

**A**. HR+HER2- – hormone receptor-positive/HER2-negative,

**B.** TNBC – triple-negative breast cancer, and

**C.** HER2+ – HER2-positive.

P values were calculated using Kruskal-Wallis (>2 groups) or Wilcoxon rank-sum (2 groups) tests. The red texts indicate statistically significant differences in the MTM/mL distributions.

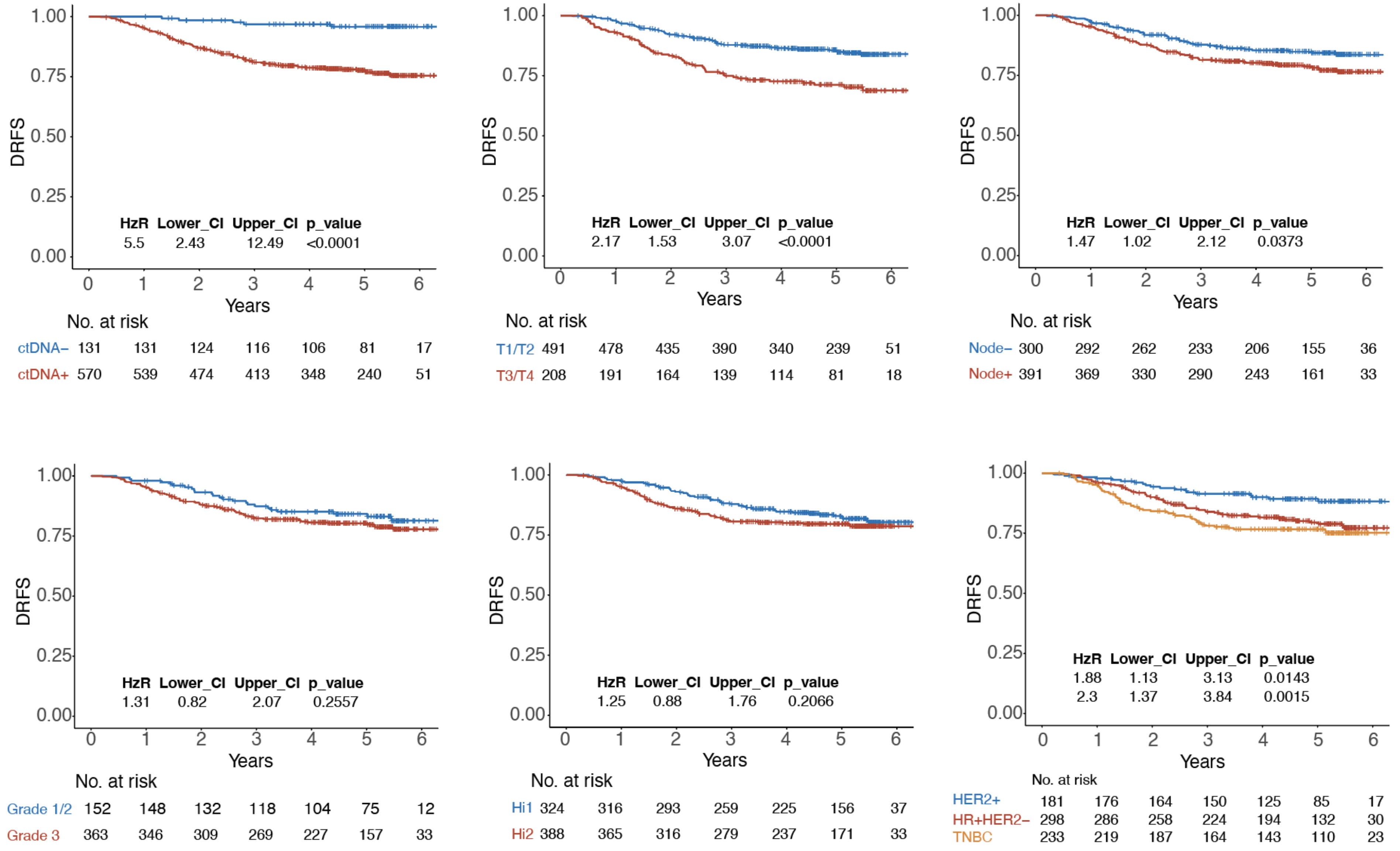
DRFS
1.00
0.75
0.50
0.25
0.00
0 1 2 3 4 5 6
Years
HzR Lower_CI Upper_CI p_value
5.5 2.43 12.49 <0.0001
No. at risk
ctDNA− 131 131 124 116 106 81 17
ctDNA+ 570 539 474 413 348 240 51
HzR Lower_CI Upper_CI p_value
2.17 1.53 3.07 <0.0001
No. at risk
T1/T2 491 478 435 390 340 239 51
T3/T4 208 191 164 139 114 81 18
HzR Lower_CI Upper_CI p_value
1.47 1.02 2.12 0.0373
No. at risk
Node− 300 292 262 233 206 155 36
Node+ 391 369 330 290 243 161 33
HzR Lower_CI Upper_CI p_value
1.31 0.82 2.07 0.2557
No. at risk
Grade 1/2 152 148 132 118 104 75 12
Grade 3 363 346 309 269 227 157 33
HzR Lower_CI Upper_CI p_value
1.25 0.88 1.76 0.2066
No. at risk
Hi1 324 316 293 259 225 156 37
Hi2 388 365 316 279 237 171 33
HzR Lower_CI Upper_CI p_value
1.88 1.13 3.13 0.0143
2.3 1.37 3.84 0.0015
No. at risk
HER2+ 181 176 164 150 125 85 17
HR+HER2− 298 286 258 224 194 132 30
TNBC 233 219 187 164 143 110 23

**Figure S5. Distant recurrence-free survival in patients grouped according to tumor characteristics at diagnosis.**

Kaplan-Meier plots compare distant recurrence-free survival (DRFS) of patients stratified according to (**top panel**) circulating tumor DNA (ctDNA) status at diagnosis (ctDNA-negative vs. ctDNA-positive), clinical T stage (T1/T2 vs. T3/T4), clinical N stage (node-negative vs. node-positive), (**bottom panel**) grade (1/2 vs. 3), MammaPrint score (high 1 vs. high 2), subtypes (HR+HER2- – hormone receptor-positive/HER2-negative, TNBC – triple-negative breast cancer, and HER2+ – HER2-positive). The hazard ratios and 95% confidence intervals (CI) from univariable Cox regression analyses are shown at the bottom of each plot. P values were calculated using the Wald test.

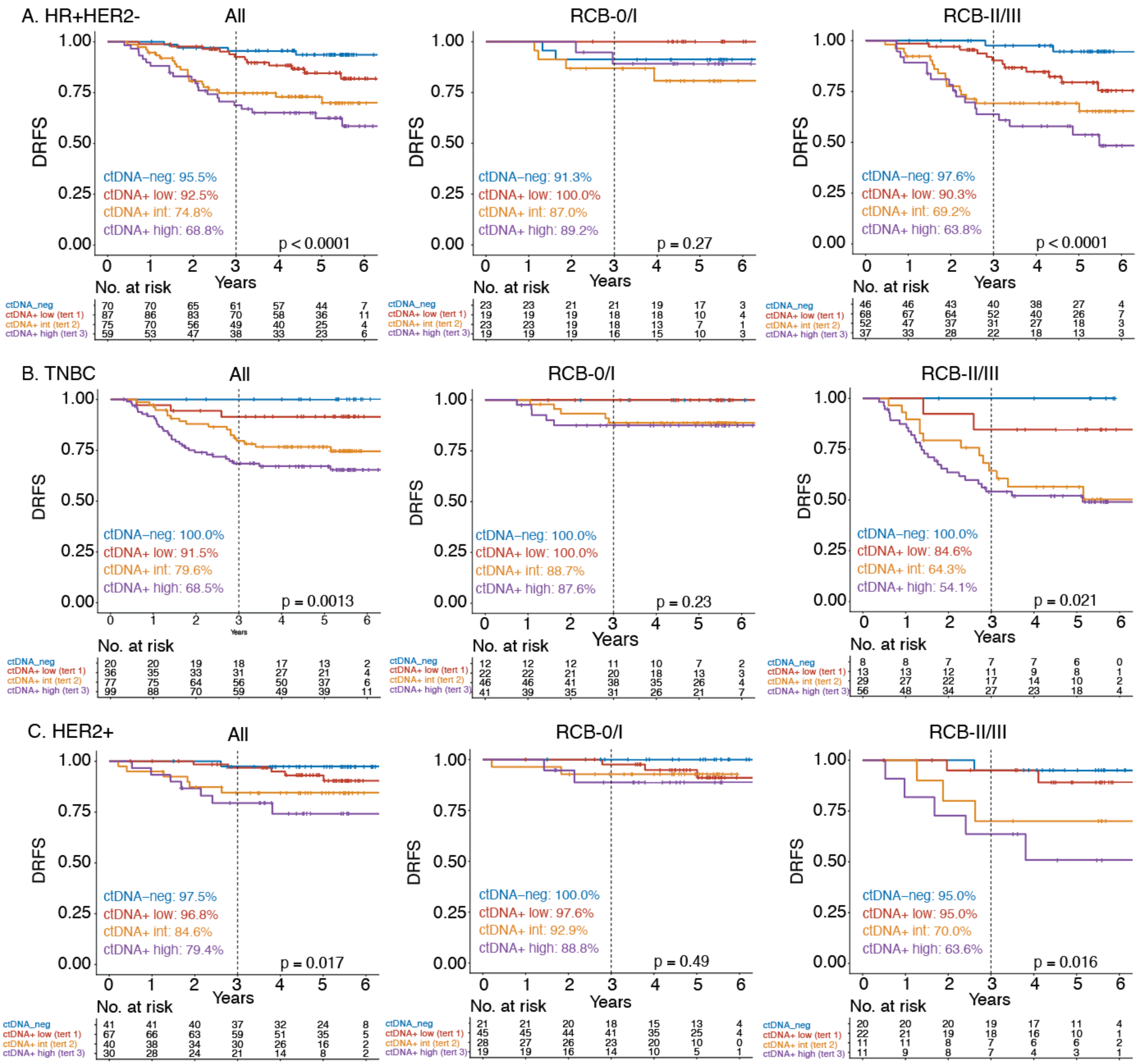

A. HR+HER2-
All
RCB-0/I
RCB-II/III
DRFS
Years
ctDNA–neg: 95.5%
ctDNA+ low: 92.5%
ctDNA+ int: 74.8%
ctDNA+ high: 68.8%
p < 0.0001
ctDNA–neg: 91.3%
ctDNA+ low: 100.0%
ctDNA+ int: 87.0%
ctDNA+ high: 89.2%
p = 0.27
ctDNA–neg: 97.6%
ctDNA+ low: 90.3%
ctDNA+ int: 69.2%
ctDNA+ high: 63.8%
p < 0.0001
No. at risk
ctDNA_neg 70 70 65 61 57 44 7
ctDNA+ low (tert 1) 87 86 83 70 58 36 11
ctDNA+ int (tert 2) 75 70 56 49 40 25 4
ctDNA+ high (tert 3) 59 53 47 38 33 23 6
ctDNA_neg 23 23 21 21 19 17 3
ctDNA+ low (tert 1) 19 19 19 18 18 10 4
ctDNA+ int (tert 2) 23 23 19 18 13 7 1
ctDNA+ high (tert 3) 19 19 19 16 15 10 3
ctDNA_neg 46 46 43 40 38 27 4
ctDNA+ low (tert 1) 68 67 64 52 40 26 7
ctDNA+ int (tert 2) 52 47 37 31 27 18 3
ctDNA+ high (tert 3) 37 33 28 22 18 13 3
B. TNBC
All
RCB-0/I
RCB-II/III
ctDNA–neg: 100.0%
ctDNA+ low: 91.5%
ctDNA+ int: 79.6%
ctDNA+ high: 68.5%
p = 0.0013
ctDNA–neg: 100.0%
ctDNA+ low: 100.0%
ctDNA+ int: 88.7%
ctDNA+ high: 87.6%
p = 0.23
ctDNA–neg: 100.0%
ctDNA+ low: 84.6%
ctDNA+ int: 64.3%
ctDNA+ high: 54.1%
p = 0.021
No. at risk
ctDNA_neg 20 20 19 18 17 13 2
ctDNA+ low (tert 1) 36 35 33 31 27 21 4
ctDNA+ int (tert 2) 77 75 64 56 50 37 6
ctDNA+ high (tert 3) 99 88 70 59 49 39 11
ctDNA_neg 12 12 12 11 10 7 2
ctDNA+ low (tert 1) 22 22 21 20 18 13 3
ctDNA+ int (tert 2) 46 46 41 38 35 26 4
ctDNA+ high (tert 3) 41 39 35 31 26 21 7
ctDNA_neg 8 8 7 7 7 6 0
ctDNA+ low (tert 1) 13 13 12 11 9 8 1
ctDNA+ int (tert 2) 29 27 22 17 14 10 2
ctDNA+ high (tert 3) 56 48 34 27 23 18 4
C. HER2+
All
RCB-0/I
RCB-II/III
ctDNA–neg: 97.5%
ctDNA+ low: 96.8%
ctDNA+ int: 84.6%
ctDNA+ high: 79.4%
p = 0.017
ctDNA–neg: 100.0%
ctDNA+ low: 97.6%
ctDNA+ int: 92.9%
ctDNA+ high: 88.8%
p = 0.49
ctDNA–neg: 95.0%
ctDNA+ low: 95.0%
ctDNA+ int: 70.0%
ctDNA+ high: 63.6%
p = 0.016
No. at risk
ctDNA_neg 41 41 40 37 32 24 8
ctDNA+ low (tert 1) 67 66 63 59 51 35 5
ctDNA+ int (tert 2) 40 38 34 30 26 16 2
ctDNA+ high (tert 3) 30 28 24 21 14 8 2
ctDNA_neg 21 21 20 18 15 13 4
ctDNA+ low (tert 1) 45 45 44 41 35 25 4
ctDNA+ int (tert 2) 28 27 26 23 20 10 0
ctDNA+ high (tert 3) 19 19 16 14 10 5 1
ctDNA_neg 20 20 20 19 17 11 4
ctDNA+ low (tert 1) 22 21 19 18 16 10 1
ctDNA+ int (tert 2) 11 11 8 7 6 6 2
ctDNA+ high (tert 3) 11 9 8 7 4 3 1

**Figure S6. ctDNA concentration at diagnosis is a prognostic factor for distant metastatic recurrence in the context of treatment response.** Kaplan-Meier plots compare distant recurrence-free survival (DRFS) among patients stratified by ctDNA concentration group [ctDNA-negative and ctDNA positive: low (tertile 1), intermediate (tertile 2), and high (tertile 3)] are shown in the leftmost column. Patients were stratified by dichotomized residual cancer burden (RCB) class. Results for patients with RCB-0 (pathologic complete response, pCR) or RCB-I (limited) are shown in the middle panel. Results for patients with RCB-II (moderate) or RCB-III (extensive) are shown in the rightmost panel. The 3-year DRFS rates are shown in the bottom-left quadrant of each plot. P values were calculated using the log-rank test. The analysis was performed across subtypes (rows):

**A**. HR+HER2- – hormone receptor-positive/HER2-negative,

**B.** TNBC – triple-negative breast cancer, and

**C.** HER2+ – HER2-positive.

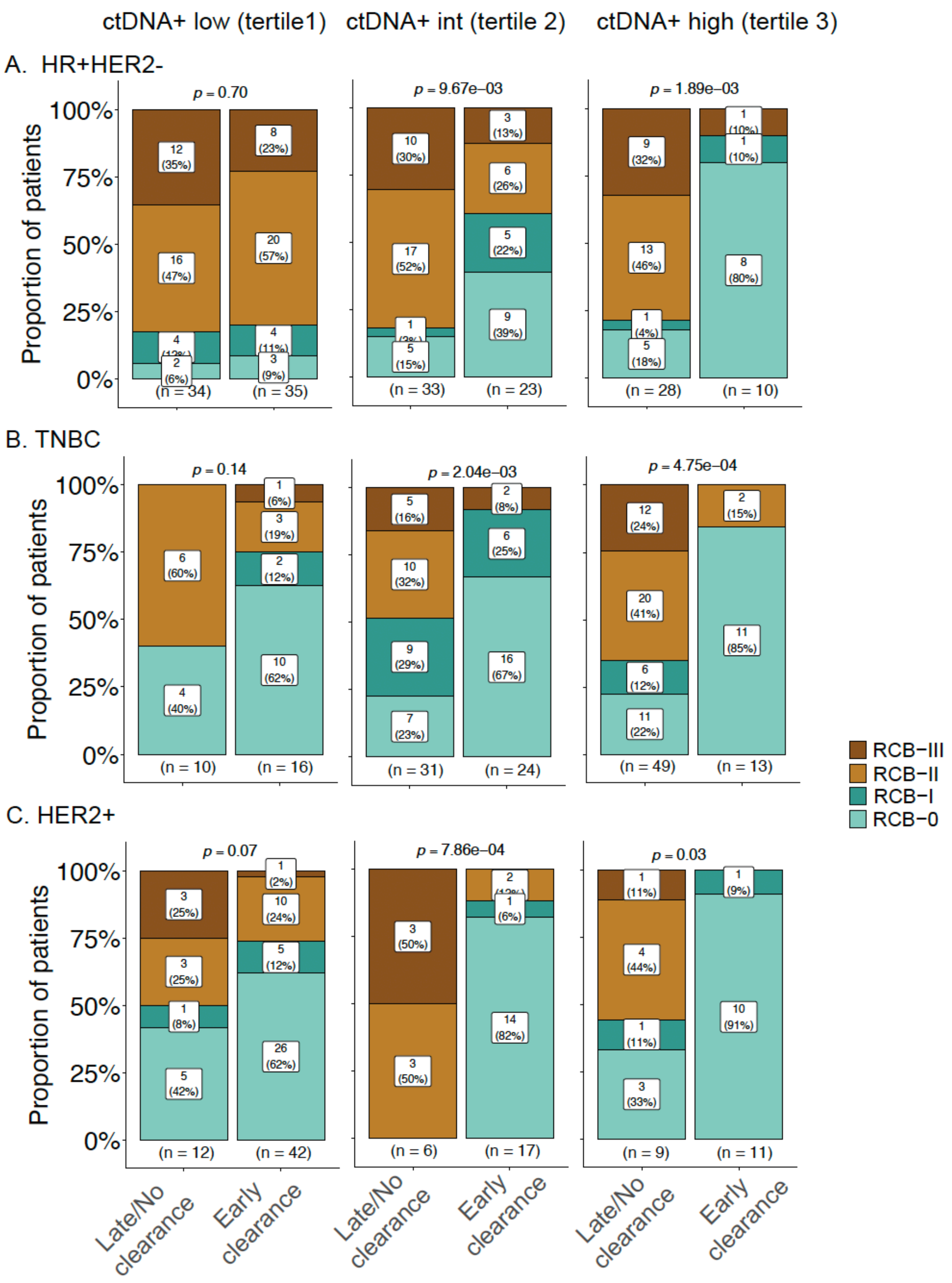
ctDNA+ low (tertile1)
ctDNA+ int (tertile 2)
ctDNA+ high (tertile 3)
A. HR+HER2-
B. TNBC
C. HER2+
Proportion of patients
p = 0.70
p = 9.67e−03
p = 1.89e−03
p = 0.14
p = 2.04e−03
p = 4.75e−04
p = 0.07
p = 7.86e−04
p = 0.03
(n = 34)
(n = 35)
(n = 33)
(n = 23)
(n = 28)
(n = 10)
(n = 10)
(n = 16)
(n = 31)
(n = 24)
(n = 49)
(n = 13)
(n = 12)
(n = 42)
(n = 6)
(n = 17)
(n = 9)
(n = 11)
Late/No clearance
Early clearance
RCB−III
RCB−II
RCB−I
RCB−0

**Figure S7. Patients across subtypes with intermediate or high ctDNA concentration at diagnosis who clear ctDNA are significantly enriched for good responders.** Patients who were ctDNA-positive at diagnosis were grouped by ctDNA concentration at diagnosis (columns): low (tertile 1), intermediate (tertile 2), and high (tertile 3). Bar plots show the proportions of patients and their residual cancer burden (RCB) class, grouped by ctDNA dynamics: Early clearance vs. Late or No clearance. 'Early clearance' included ctDNA-positive patients whose ctDNA cleared at week 3 after treatment initiation (T1). 'Late or no clearance' included those whose ctDNA cleared at week 12 (T2), or post-neoadjuvant therapy (T3), or who had no clearance at T3. P values were calculated using the Chi-squared test. The analysis was performed across subtypes (rows):

**A**. HR+HER2- – hormone receptor-positive/HER2-negative,

**B.** TNBC – triple-negative breast cancer, and

**C.** HER2+ – HER2-positive.

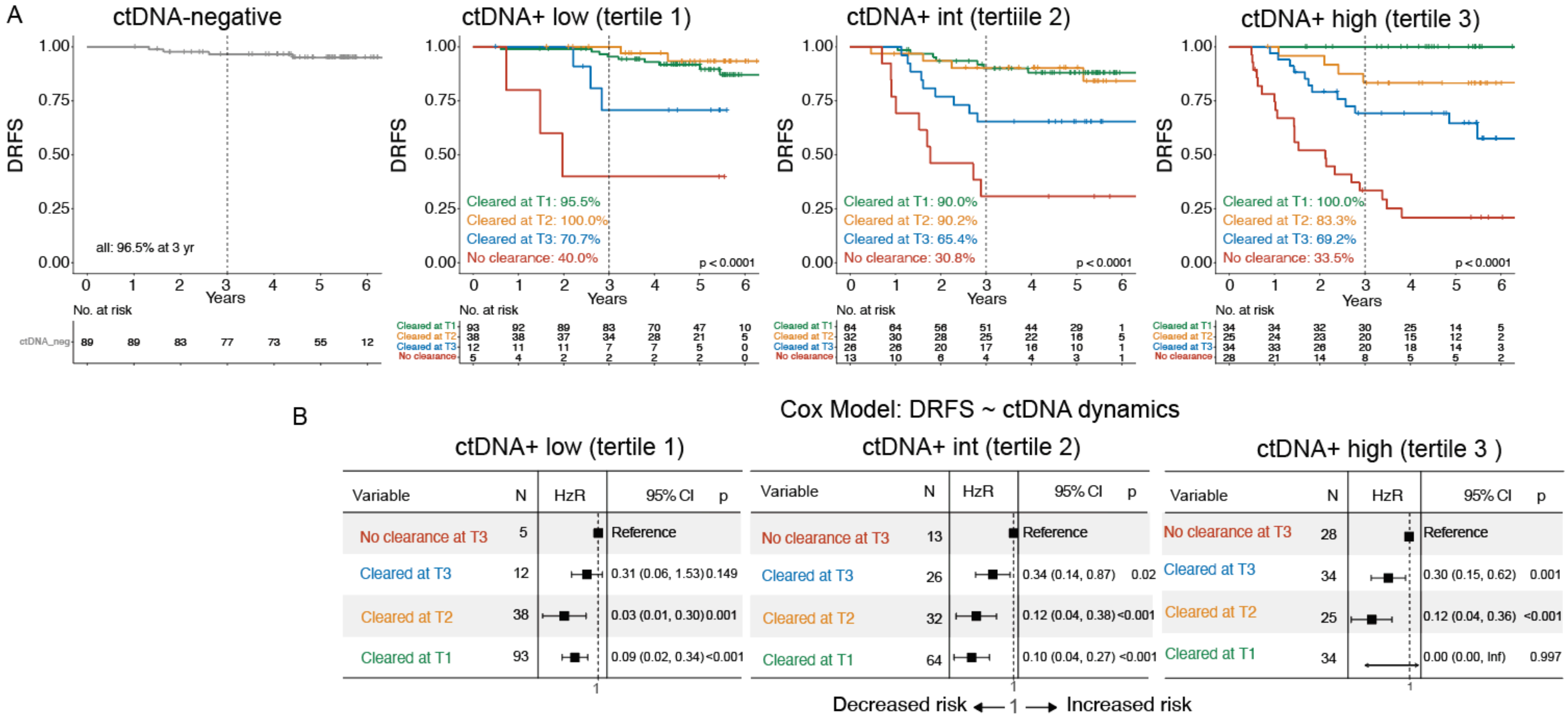

**Figure S8. Correlation between ctDNA clearance and risk of metastatic recurrence across subtypes, stratified by ctDNA concentration at diagnosis.**

**A.** Kaplan-Meier plots compare distant recurrence-free survival (DRFS) across 4 patient groups by ctDNA concentration at diagnosis [ctDNA-negative and ctDNA-positive: low (tertile 1), intermediate (tertile 2), and high (tertile 3)]. Each ctDNA concentration group was stratified by ctDNA dynamics: persistent negative, cleared at week 3 (T1) or week 12 (T2) after treatment initiation, cleared at post-NAT before surgery (T3), and no clearance at T3. The 3-year DRFS rates according to ctDNA dynamics are shown in the lower-left quadrant. The P value was calculated using the log-rank test. The same analysis was performed across HR/HER2 subtypes (**Figure S9**) and in patients with residual cancer burden: RCB-II (moderate) or RCB-III (extensive) (**Figure 5**).

**B.** Cox regression analysis was performed to estimate hazard ratios and 95% confidence intervals (CI) for ctDNA-positive patients grouped by ctDNA dynamics: cleared at T1, T2, or T3, and no clearance at T3. The no clearance group served as the reference.

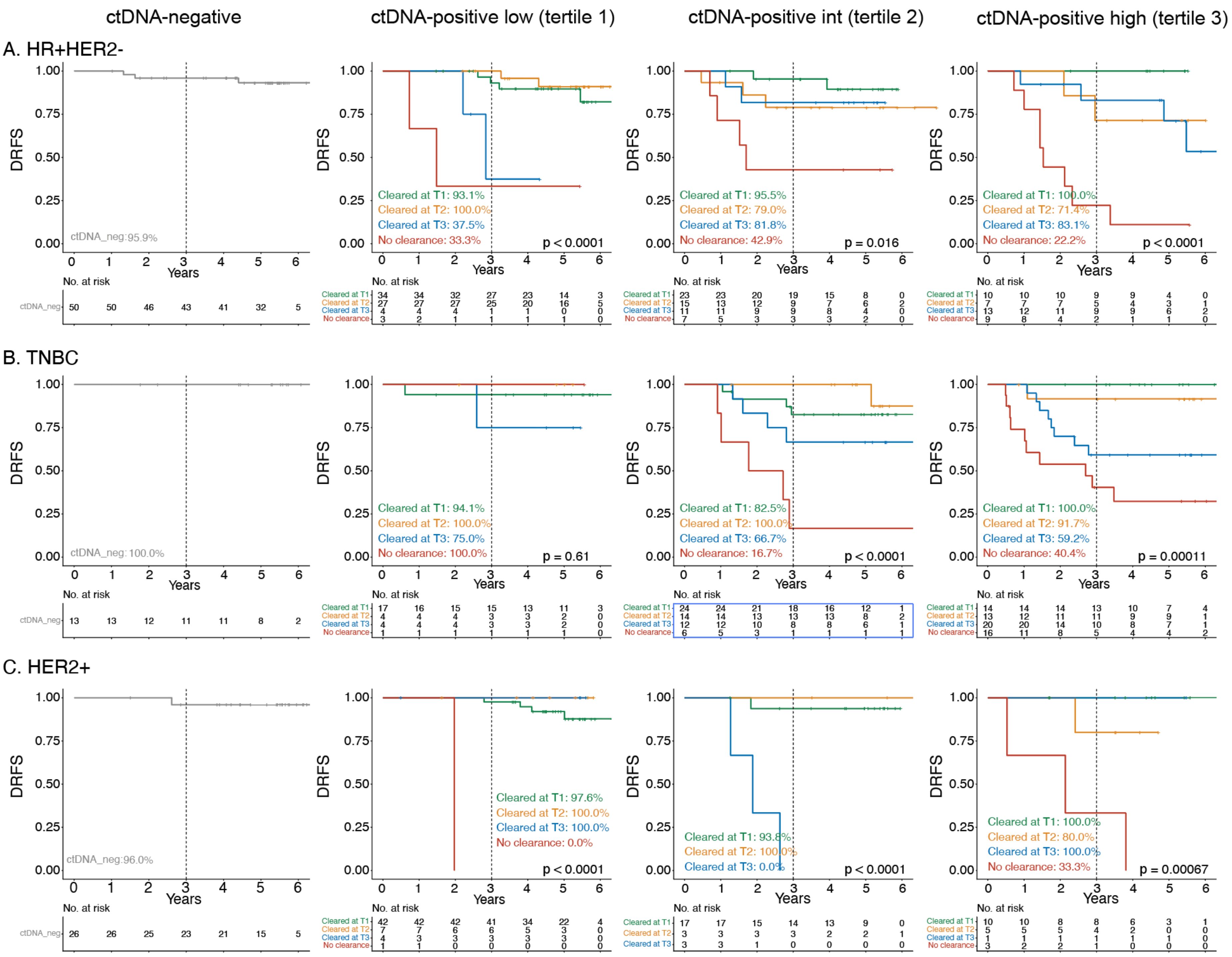
ctDNA-negative
ctDNA-positive low (tertile 1)
ctDNA-positive int (tertile 2)
ctDNA-positive high (tertile 3)
A. HR+HER2-
ctDNA_neg:95.9%
Cleared at T1: 93.1%
Cleared at T2: 100.0%
Cleared at T3: 37.5%
No clearance: 33.3%
p < 0.0001
Cleared at T1: 95.5%
Cleared at T2: 79.0%
Cleared at T3: 81.8%
No clearance: 42.9%
p = 0.016
Cleared at T1: 100.0%
Cleared at T2: 71.4%
Cleared at T3: 83.1%
No clearance: 22.2%
p < 0.0001
B. TNBC
ctDNA_neg: 100.0%
Cleared at T1: 94.1%
Cleared at T2: 100.0%
Cleared at T3: 75.0%
No clearance: 100.0%
p = 0.61
Cleared at T1: 82.5%
Cleared at T2: 100.0%
Cleared at T3: 66.7%
No clearance: 16.7%
p < 0.0001
Cleared at T1: 100.0%
Cleared at T2: 91.7%
Cleared at T3: 59.2%
No clearance: 40.4%
p = 0.00011
C. HER2+
ctDNA_neg:96.0%
Cleared at T1: 97.6%
Cleared at T2: 100.0%
Cleared at T3: 100.0%
No clearance: 0.0%
p < 0.0001
Cleared at T1: 93.8%
Cleared at T2: 100.0%
Cleared at T3: 0.0%
p < 0.0001
Cleared at T1: 100.0%
Cleared at T2: 80.0%
Cleared at T3: 100.0%
No clearance: 33.3%
p = 0.00067
DRFS
Years
No. at risk

**Figure S9. Correlation between ctDNA clearance and risk of metastatic recurrence in all patients (complete ctDNA data) stratified by ctDNA concentration at diagnosis.** Kaplan-Meier plots compare distant recurrence-free survival (DRFS) across 4 patient groups, stratified by ctDNA concentration at diagnosis (columns): ctDNA-negative and ctDNA-positive: low (tertile 1), intermediate (tertile 2), and high (tertile 3). Each ctDNA concentration group was stratified by ctDNA dynamics: persistent negative, cleared at week 3 (T1) or week 12 (T2) after treatment initiation, cleared at post-NAT before surgery (T3), and no clearance at T3. The 3-year DRFS rates, according to ctDNA dynamics, are shown in the lower-left quadrant. The P value was calculated using the log-rank test. The analysis was performed across subtypes (rows):

**A**. HR+HER2- – hormone receptor-positive/HER2-negative,

**B.** TNBC – triple-negative breast cancer, and

**C.** HER2+ – HER2-positive.

**Table S1. Patient and tumor characteristics.** Patients were grouped into ctDNA-negative and ctDNA-positive at diagnosis. Individuals who tested positive for ctDNA were subsequently divided into 3 equally sized groups based on ctDNA concentration: low (tertile 1), intermediate (tertile 2), and high (tertile 3). For categorical variables, proportions were compared using the Chi-squared test; for the continuous variable (Age at diagnosis), distributions were compared using a t-test. This study includes 712 patients from the 723-patient cohort previously described in detail [3]. Abbreviations: HR – hormone receptor; RCB – residual cancer burden; SD – standard deviation; TNBC – triple negative breast cancer.

| **Clinicopathologic variable** | **ctDNA groups at diagnosis** | **ctDNA-negative** | **ctDNA+ low (tertile 1)** | **ctDNA+ int (tertile 2)** | **ctDNA+ high (tertile 3)** | **Missing ctDNA at diagnosis** | **Total N**** | **p** |
|---|---|---|---|---|---|---|---|---|
| | | n=133 | n=193 | n=193 | n=193 | n=11 | n=723 | |
| | | n (%) | n (%) | n (%) | n (%) | n (%) | n (%) | |
| Subtype | HR+HER2- | 71 (53.4) | 88 (45.6) | 75 (38.9) | 59 (30.6) | 7 (63.6) | 300 (41.5) | <0.001 |
| | TNBC | 20 (15.0) | 37 (19.2) | 78 (40.4) | 101 (52.3) | 1 (9.1) | 237 (32.8) | |
| | HER2+ | 42 (31.6) | 68 (35.2) | 40 (20.7) | 33 (17.1) | 3 (27.3) | 186 (25.7) | |
| | Missing | 0 (0.0) | 0 (0.0) | 0 (0.0) | 0 (0.0) | 0 (0.0) | 0 (0.0) | |
| Clinical T Stage | T1/T2 | 111 (83.5) | 142 (73.6) | 138 (71.5) | 98 (50.8) | 7 (63.6) | 496 (68.6) | <0.001 |
| | T3/T4 | 20 (15.0) | 49 (25.4) | 51 (26.4) | 90 (46.6) | 4 (36.4) | 214 (29.6) | |
| | Missing | 2 (1.5) | 2 (1.0) | 4 (2.1) | 5 (2.6) | 0 (0.0) | 13 (1.8) | |
| Clinical N Stage | Node-positive | 53 (39.8) | 100 (51.8) | 109 (56.5) | 130 (67.4) | 9 (81.8) | 401 (55.5) | <0.001 |
| | Node-negative | 77 (57.9) | 88 (45.6) | 77 (39.9) | 57 (29.5) | 2 (18.2) | 301 (41.6) | |
| | Missing | 3 (2.3) | 5 (2.6) | 7 (3.6) | 6 (3.1) | 0 (0.0) | 21 (2.9) | |
| Grade | 1/2 | 59 (44.4) | 52 (26.9) | 25 (13.0) | 13 (6.7) | 4 (36.4) | 153 (21.2) | <0.001 |

| | | | | | | | | |
|---|---|---|---|---|---|---|---|---|
| | 3 | 44 (33.1) | 83 (43.0) | 119 (61.7) | 122 (63.2) | 3 (27.3) | 371 (51.3) | |
| | Missing | 30 (22.6) | 58 (30.1) | 49 (25.4) | 58 (30.1) | 4 (36.4) | 199 (27.5) | |
| MammaPrint | High 1 | 101 (75.9) | 117 (60.6) | 62 (32.1) | 39 (20.2) | 9 (81.8) | 328 (45.4) | <0.001 |
| | High 2 | 32 (24.1) | 76 (39.4) | 131 (67.9) | 154 (79.8) | 2 (18.2) | 395 (54.6) | |
| | Missing | 0 (0.0) | 0 (0.0) | 0 (0.0) | 0 (0.0) | 0 (0.0) | 0 (0.0) | |
| Residual Cancer Burden (RCB) | RCB-0 | 38 (28.6) | 67 (34.7) | 70 (36.3) | 66 (34.2) | 0 (0.0) | 241 (33.3) | 0.16 |
| | RCB-I | 18 (13.5) | 19 (9.8) | 27 (14.0) | 14 (7.3) | 2 (18.2) | 80 (11.1) | |
| | RCB-II | 54 (40.6) | 73 (37.8) | 58 (30.1) | 65 (33.7) | 5 (45.5) | 255 (35.3) | |
| | RCB-III | 21 (15.8) | 32 (16.6) | 34 (17.6) | 41 (21.2) | 4 (36.4) | 132 (18.3) | |
| | Missing | 2 (1.5) | 2 (1.0) | 4 (2.1) | 7 (3.6) | 0 (0.0) | 15 (2.1) | |
| Age | Mean (SD) | 49.0 (10.3) | 48.0 (11.0) | 50.5 (10.9) | 48.2 (11.3) | 46.1 (12.0) | 48.9 (11.0) | 0.14 |
| | Missing | 0 (0.0) | 0 (0.0) | 0 (0.0) | 0 (0.0) | 0 (0.0) | 0 (0.0) | |

**Total N from Magbanua et al [3]

**Table S2. Treatment received in the I-SPY2 trial.** Patients were randomized to treatment arms. ctDNA data at diagnosis were available for 712 patients.

| Treatment | No. of patient s |
|---|---|
| Paclitaxel | 159 |
| Paclitaxel + Pertuzumab +/- Trastuzumab | 117 |
| Paclitaxel + ABT 888 + Carboplatin | 58 |
| Paclitaxel + PD-1 inhibitor 8 cycles | 56 |
| Paclitaxel + MK-2206 +/- Trastuzumab | 53 |
| Paclitaxel + Durvalumab + Olaparib | 52 |
| Irinotecan + Talazoparib | 46 |
| Paclitaxel + PD-1 inhibitor 4 cycles | 44 |
| T-DM1 + Pertuzumab | 42 |
| SD-101 +PD-1 inhibitor | 17 |
| Paclitaxel + Ganetespib | 16 |
| Paclitaxel + Ganitumab | 15 |
| SGN-LIV1A | 11 |
| Paclitaxel + AMG 386 +/- Trastuzumab | 10 |
| Paclitaxel + Neratinib | 6 |
| Paclitaxel + Cemiplimab +/- REGN3767 | 5 |
| Paclitaxel + Pertuzumab + Trastuzumab + Tucatinib | 3 |
| Paclitaxel + Patritumab + Trastuzumab | 2 |

**Table S3. Patients with complete ctDNA data at all four time points.** The table shows the number of patients included in analyses of ctDNA dynamics (timing of clearance). ctDNA testing was performed on blood samples collected at diagnosis (pretreatment, T0), at week 3 (T1), at week 12 (T2) after treatment initiation, and post-NAT before surgery (T3). Abbreviations: HR – hormone receptor; TNBC – triple negative breast cancer.

| **ctDNA dynamics** | **ctDNA status at each time point (T0/T1/T2/T3)** | **HR+HER2-** | | **TNBC** | | **HER2+** | |
|---|---|---|---|---|---|---|---|
| | | **n** | **%** | **n** | **%** | **n** | **%** |
| Persistent negative | ctDNA-/-/-/- | 50 | 23.4 | 13 | 8.2 | 26 | 21.1 |
| Cleared at week 3 (T1) | ctDNA+/-/-/- | 68 | 31.8 | 55 | 34.6 | 70 | 56.9 |
| Cleared at week 12 (T2) | ctDNA+/+/-/- | 49 | 22.9 | 31 | 19.5 | 15 | 12.2 |
| Cleared post-NAT before surgery (T3) | ctDNA+/+/+/- or ctDNA+/-/+/- | 28 | 13.1 | 37 | 23.3 | 8 | 6.5 |
| No clearance at T3 | ctDNA+/+/+/+, ctDNA+/-/+/+, or ctDNA+/+/-/+ | 19 | 8.9 | 23 | 14.5 | 4 | 3.3 |
| | Total | 214 | 100. | 159 | 100. | 123 | 100 |
