## Supplementary Material for "Circulating tumor DNA concentration at diagnosis is a modifiable prognostic factor for distant metastatic recurrence in patients with high-risk breast cancer receiving neoadjuvant therapy"

The following IRB approved the clinical research protocol.

| Columbia NYC | Columbia Research IRB |
| --- | --- |
| Emory University | Emory IRB |
| Georgetown University | MedStar Health Research Institute |
| Inova Health System | Inova Health System IRB |
| Loyola University Medical Center | Loyola University Chicago Health Sciences Division IRB |
| Mayo Clinic, Rochester - MN | Mayo Clinic IRB |
| Mayo Clinic, Scottsdale - AZ | Mayo Clinic IRB |
| Moffitt Cancer Center | *Chesapeake (Liberty) IRB, after A19 it was Advarra IRB |
| Oregon Health and Science University | OHSU IRB |
| Rutgers Cancer Institute of New Jersey | Rutgers/RWJMS IRB |
| Sanford | Sanford Health IRB |
| Swedish Cancer Institute | Western IRB (WIRB) |
| The Ohio State University | The Office of Responsible Research Practices (ORRP) |
| UC Davis Health | UC Davis IRB |
| University of Alabama Birmingham | UAB IRB |
| University of Arizona | University of Arizona IRB |
| University of California San Diego | UCSD IRB (Human Rights Protection Program) |
| University of California San Francisco | UCSF IRB (Human Subject Protection Program) |
| University of Chicago - Main | AURA IRB |
| University of Colorado - Main | Colorado Multiple (COM) IRB |
| University of Kansas | KUMC Human Research Protection Program |
| University of Minnesota | University of Minnesota IRB |
| University of Pennsylvania | UPENN IRB |
| University of Rochester Medical Center (URMC) | Research Subjects Review Board (RSRB) |
| University of Southern California | USC Health Sciences Campus IRB |
| University of Texas MD Anderson | UT MD Anderson Cancer Center IRB |
| University of Texas Southwestern | UTSW Center eIRB System |
| University of Utah (Huntsman Cancer Institute) | University of Utah IRB |
| University of Washington | Fred Hutchinson Cancer Research Ctr IRB |
| Vanderbilt | Vanderbilt IRB |
| Wake Forest | Wake Forest University Health Sciences IRB |
| Yale | Yale IRB |

*Liberty IRB (also known as Schulman IRB) merged with Chesapeake IRB and then became Advarra IRB in 2017
